# Gene-Excessive Sleepiness Interactions Suggest Treatment Targets for Obstructive Sleep Apnea Subtype

**DOI:** 10.1101/2024.10.25.24316158

**Authors:** Pavithra Nagarajan, Nuzulul Kurniansyah, Jiwon Lee, Sina A. Gharib, Yushan Xu, Yiyan Zhang, Brian Spitzer, Tariq Faquih, Hufeng Zhou, Eric Boerwinkle, Han Chen, Daniel J. Gottlieb, Xiuqing Guo, Nancy L. Heard-Costa, Bertha A. Hidalgo, Daniel Levy, Peter Y. Liu, Hao Mei, Rebecca Montalvan, Sutapa Mukherjee, Kari E. North, George T. O’Conner, Lyle J. Palmer, Sanjay R. Patel, Bruce M. Psaty, Shaun M. Purcell, Laura M. Raffield, Stephen S. Rich, Jerome I. Rotter, Richa Saxena, Albert V Smith, Katie L. Stone, Xiaofeng Zhu, TOPMed Sleep Trait WG, Brian E. Cade, Tamar Sofer, Susan Redline, Heming Wang

## Abstract

Obstructive sleep apnea (OSA) is a multifactorial sleep disorder characterized by a strong genetic basis. Excessive daytime sleepiness (EDS) is a symptom that is reported by a subset of OSA patients, persisting even after treatment with continuous positive airway pressure (CPAP). It is recognized as a clinical subtype underlying OSA carrying alarming heightened cardiovascular risk. Thus, conceptualizing EDS as an exposure variable, we sought to investigate EDS’s influence on genetic variation linked to apnea-hypopnea index (AHI), a diagnostic measure of OSA severity. This study serves as the first large-scale genome-wide gene x environment interaction analysis for AHI, investigating the interplay between its genetic markers and EDS across and within specific sex. Our work pools together whole genome sequencing data from seven cohorts, enabling a diverse dataset (four population backgrounds) of over 11,500 samples. Among the total 16 discovered genetic targets with interaction evidence with EDS, eight are previously unreported for OSA, including *CCDC3*, *MARCHF1*, and *MED31* identified in all sexes; *TMEM26*, *CPSF4L*, and *PI4K2B* identified in males; and *RAP1GAP* and *YY1* identified in females. We discuss connections to insulin resistance, thiamine deficiency, and resveratrol use that may be worthy of therapeutic consideration for excessively sleepy OSA patients.

## INTRODUCTION

Obstructive sleep apnea (OSA) is characterized by recurrent upper airway collapse and obstruction, resulting in arousal and oxygen desaturation events that induce variable degrees of sympathetic activation, inflammation, endothelial dysfunction, and metabolic perturbations ^1^. The clinical presentation of this disorder can vary and includes both disturbed sleep (insomnia) and excessive daytime sleepiness (EDS) ^1^. A strong genetic basis has been established for OSA with heritability estimates between 69% to 83% in twin studies, 25% to 40% in family studies, and up to 21% from population-based genome-wide association studies (GWAS) ^2,3^. Adequately characterizing the genetic architecture of OSA can provide insight into disease mechanisms and better therapeutic regimens, ultimately improving patient outcomes.

In OSA, EDS is found in over 30% of patients ^4^. Although often exhibiting improvement with OSA treatment, EDS persists in 9% to 22% of CPAP-adherent individuals ^4,5^. Patients with EDS (excessive sleepy OSA subtype) are reported to experience higher risk of incident cardiovascular disease, potentially reflecting elevated inflammation and a more severe endophenotype (high airway collapsibility, low arousal threshold) ^1,6,7^. EDS can reflect behavioral factors associated with OSA risk such as insufficient sleep duration, poor diet quality, and reduced physical activity ^8–12^. As a marker of underlying physiological conditions and environmental exposures, EDS may moderate genetic effects that influence severity of OSA and its cardiovascular risk, with possible pathways linked to the microbiome, systemic inflammation, and adipose tissue function ^1,13^.

We thus sought to investigate whether there is evidence of interaction between EDS and genetic variant effects for apnea-hypopnea index (AHI). We conducted genome-wide interaction analyses with single common variant effects, and rare variant gene set-based effects, in over 11,500 individuals using National Heart, Lung and Blood Institute Trans-Omics for Precision Medicine (TOPMed) data ^14^. A dataset comprising of multiple race/ethnicities (African American/Black [AFR], Asian [ASN], Caucasian/White/European [EUR], Hispanic/Latino [HIS]) was consolidated for this analysis with stratification according to sex (males, females, all sex). Preliminary results have been previously reported in the form of an abstract ^15^.

## RESULTS

### Common Variant Analysis

Common variant analysis with whole genome sequencing (WGS) data in combined sex samples revealed two intronic variants with strong interaction with EDS - *rs281851* (P_GxE:_ 6.59e-09, P_G,GxE:_ 2.2e-08) mapped to *CCDC3*, and *rs13118183* (P_GxE_: 2.92e-08, P_G,GxE:_ 4.7e-08) mapped to *MARCHF1* (Table 1, Figure 2A). Variant to gene mapping by FUMA SNP2GENE (using position, expression quantitative trait loci, chromatin interaction methods) additionally identified *UPF2, DHTKD1, SEC61A2, NUDT5, CDC123, CAMK1D, OPTN*, and *PHYH* mapped to *rs281851* and *NAF1, NPY1R, NPY5R, TKTL2, FAM218A,* and *TRIM60* mapped to *rs13118183* (Table 1, Supplementary Table 2).

**Table 1.** Genetic Variants Interacting with EDS Identified in Common Variant Discovery Analysis.

| *RSID* * | Chr:Position<br>(b38) | Effect<br>Allele/<br>Alternative<br>Allele | Effect Allele<br>Frequency | Gene Locus | N | MAIN EFFECT |  | INTERACTION<br>EFFECT |  | JOINT<br>EFFECT |
| --- | --- | --- | --- | --- | --- | --- | --- | --- | --- | --- |
|  |  |  |  |  |  | Beta<br>(Standard<br>Error) | P-Value | Beta<br>(Standard<br>Error) | P-Value | P-Value |
| ***RS13118183*** | 4:164200775 | A/G | 0.94 | *NAF1*,<br>*NPY1R*,<br>*NPY5R*,<br>*TKTL2*,<br>*MARCHF1*,<br>*FAM218A*,<br>*TRIM60* | 11614 | -0.0094<br>(0.0025) | 1.50e-04 | 0.034<br>(0.0062) | 2.92e-08 | 4.70e-08 |
| ***RS281851*** | 10:12924495 | A/C | 0.59 | *UPF2*,<br>*DHTKD1*,<br>*SEC61A2*,<br>*NUDT5*,<br>*CDC123*,<br>*CAMK1D*,<br>*CCDC3*,<br>*OPTN*,<br>*PHYH* | 11615 | -0.0010<br>(0.0011) | 3.82e-01 | 0.0187<br>(0.0032) | 6.59e-09 | 2.23e-08 |

\*These significant variants (RSID) passed genome-wide significance criteria ( $p < 5e-08$ ) for the 1df GxE interaction test and 2df G,GxE joint test, and were identified in combined sex analysis.

### Rare Variant Set-Based Analysis

Rare variant set-based analysis with WGS data identified *SCUBE2* in combined sex, and *TMEM26* and *CPSF4L* in male sex (P_G,GxE_<3e-06, P_GxE_<P_G_) (Table 2). Meta-analysis of WGS and imputed genotype data identified *UBLCP1* and *MED31* in combined sex; *YY1, CPNE5, MYMX, ZNF773, YBEY*, and *RAP1GAP* in female sex; and *PI4K2B, IQCB1,* and *CORO1A* in male sex (Table 3) (P_G,GxE_<3e-06, P_GxE_<P_G_). Among these, *YY1* additionally showed significance in the GxE interaction effect (P_GxE_<3e-06) (Table 3).

**Table 2.** Genes Interacting with EDS Identified in Rare Variant Set-Based Discovery Analysis.

| *Gene\** | *Position* | *Sex* | *Dataset* | *Number of Variants* | *Main Effect P-Value* | *Interaction Effect P-Value* | *Joint Effect P-Value* |
| --- | --- | --- | --- | --- | --- | --- | --- |
| ***SCUBE2*** | 11:9,019,476-9,138,114 | Combined | WGS | 25 | 4.16e-02 | 2.02e-05 | 3.12e-06 |
|  |  |  | Imputed | 3 | 4.97e-01 | 7.91e-01 | 7.60e-01 |
| ***TMEM26<sup>†</sup>*** | 10:61,406,642-61,453,381 | Male | WGS | 1 | 9.63e-03 | 3.63e-05 | 2.54e-06 |
| ***CPSF4L*** | 17:73,248,449-73,262,352 | Male | WGS | 5 | 9.19e-05 | 1.69e-05 | 2.39e-08 |
|  |  |  | Imputed | 1 | 5.71e-01 | 9.66e-01 | 8.80e-01 |

\*These genes passed Bonferroni-corrected significance criteria ( $p < 3e-06$ ) for the joint G,GxE test with stronger interaction signal relative to the main genetic effect ( $P_G > P_{G \times E}$ ). <sup>†</sup>Variant sets mapped to TMEM26 were not available in imputed genotype data.

**Table 3.** Genes Interacting with EDS Identified in Rare Variant Set-Based Meta-Analysis.

| *Gene* * | *Position* | *Sex* | *Number of Variants* | *Main Effect P-Value* | *Interaction Effect P-Value* | *Joint Effect P-Value* |
| --- | --- | --- | --- | --- | --- | --- |
| **UBLCP1** | 5:159,263,290-159,286,036 | Combined | 2 | 4.75e-04 | 2.29e-05 | 1.96e-07 |
| **MED31** | 17:6,643,311-6,651,634 | Combined | 3 | 1.51e-02 | 4.49e-06 | 3.22e-07 |
| **RAP1GAP** | 1:21,596,221-21,669,357 | Female | 2 | 1.73e-01 | 7.48e-06 | 2.37e-06 |
| **CPNE5** | 6:36,740,775-36,839,444 | Female | 9 | 3.30e-04 | 2.07e-04 | 1.08e-06 |
| **MYMX** | 6:44,216,926-44,218,234 | Female | 1 | 9.13e-04 | 4.15e-05 | 3.64e-07 |
| **YY1** | 14:100,238,298-100,282,788 | Female | 1 | 2.36e-01 | 2.13e-06 | 7.08e-07 |
| **ZNF773** | 19:57,499,915-57,518,404 | Female | 6 | 5.46e-04 | 2.51e-05 | 1.41e-07 |
| **YBEY** | 21:46,286,342-46,297,751 | Female | 3 | 8.62e-02 | 1.16e-05 | 2.35e-06 |
| **IQCB1** | 3:121,769,761-121,835,079 | Male | 9 | 7.69e-02 | 7.48e-06 | 2.60e-06 |
| **PI4K2B** | 4:25,160,663-25,279,204 | Male | 4 | 3.50e-02 | 3.54e-06 | 6.28e-07 |
| **CORO1A** | 16:30,182,827-30,189,076 | Male | 2 | 7.96e-03 | 1.02e-05 | 6.07e-07 |

\*These genes passed Bonferroni-corrected significance criteria ( $P < 3e-06$ ) for the GxE interaction test or for the joint G,GxE test with stronger interaction signal relative to the main genetic effect ( $P_G > P_{G \times E}$ ).

### Validation of Results

For WGS common variant results (*rs281851*, *rs13118183*) imputed genotype samples did not contain either variant. For rare variant WGS gene-set analysis results (*SCUBE2, TMEM26, CPSF4L*) imputed genotype samples did not replicate findings, showing different variant sets available for each gene compared to WGS data (Supplementary Table 3).

### Secondary Findings

Genetic associations with AHI identified by the main genetic effect B_G_ (not considering role of EDS interaction) in the WGS discovery dataset are reported in Supplementary Tables 4-6.

### Unreported Gene Targets for OSA

The variants from common variant analysis and genes from rare variant set-based analysis were assessed with prior OSA trait-related GWAS and three catalogs: GWAS Catalog, PheWeb, Sleep Disorders Knowledge Portal ^3,16–18^ (Supplementary Table 7). *SCUBE2, UBLCP1, CPNE5, MYMX, ZNF773, YBEY, IQCB1,* and *CORO1A* are identified in a prior gene-based analysis for sleep-disordered breathing traits with samples overlapping this GxE work^16^. *SCUBE2* is additionally reported in the Sleep Disorders Knowledge Portal for sleep apnea syndrome (P: 3.7e-04; common variants). *rs281851* (intronic variant of *CCDC3*)*, rs13118183* (intronic variant of *MARCHF1*)*, TMEM26, CPSF4L, MED31, YY1, RAP1GAP*, and *PI4K2B* are previously unreported for OSA (Supplementary Table 7). Four of these have been reported for cardiometabolic traits including *TMEM26* for PR interval, hypertension, systolic blood pressure, and diastolic blood pressure; *MED31* for systolic blood pressure; *RAP1GAP* for abdominal aortic calcification levels; and *YY1* for aortic atherosclerosis, and pulse pressure.

### Effects in EDS vs non-EDS groups

We performed EDS-stratified analysis for the significant interaction loci (*rs281851*, *rs13118183*) to compare how the effect estimate (B_G_) changes dependent on whether an individual experiences excessive daytime sleepiness. Figure 2B and C and Supplementary Table 8 show that for individuals who have the allele A for rs281851 or allele A for rs13118183, the presence of EDS increases AHI. For rare variant set-based analysis differences in the variants mapped to each gene in each EDS group was observed (Supplementary Table 9).

### Bioinformatics Analysis

MAGMA analysis run on FUMA’s SNP2GENE platform with WGS common variant GxE interaction summary statistics revealed tissue enrichment in breast mammary tissue and tibial nerve for females (Supplementary Table 10). MAGMA gene-based analysis identified *NOP53, EYA2*, and *ZNF563* in combined sex and *WDR19* in females (Supplementary Table 11).

Open Targets Platform mouse model data identified functional roles in immune system response (*MARCHF1, EYA2, RAP1GAP, CPNE5, CORO1A*), adipose tissue (*CCDC3, IQCB1*), metabolism (*CCDC3, RAP1GAP, CPSF4L*), nervous system or behavior (*UBLCP1, MED31, EYA2, CPNE5, YY1, IQCB1, CORO1A, WDR19*), respiratory system (*MYMX, YY1*), cardiovascular system (*IQCB1*), and craniofacial measures (*WDR19, MED31*) (Supplementary Table 12). Open Targets Genetics revealed associations to medication use for lung disease (atrovent-*YY1*, ventolin-*WDR19*), hypertension or cardiovascular disease (bendroflumethiazide-*CORO1A*, atenolol-*TMEM26,* atenolol-*MARCHF1*, bumetanide-*MYMX*, dipyridamole-*TMEM26*), and cholesterol (statin-*NOP53*, statin-*MYMX*) (Supplementary Data).

STRINGdb identified enriched terms (FDR<0.05) from the resultant protein-protein interaction network built by the *rs281851* and *rs13118183* gene loci (Supplementary Table 13). Enriched gene ontology (GO) Molecular Function terms related to the highly conserved pancreatic polypeptide hormone family *NPY-PYY-PP* (*NPY1R, NPY5R*) and thiamine pyrophosphate-transketolase activity (*DHTKD1, TKTL2*) (Supplementary Table 13).

DGIdb revealed the following drug-gene interactions: velneperit (prior Phase II clinical trial for obesity) for *NPY5R* and losartan (FDA-approved for hypertension) for *CAMK1D* (Supplementary Table 14).

Qiagen’s Ingenuity Pathway Analysis software constructed a fully-connected interactome with the primary 16 genes from Tables 1-3 provided as input (Supplementary Fig. 1). The identified connections included neuroinflammation and nerve-function (*TRIM67, HTT*), inflammation (*TNF, INFG*), tumor suppressor (*TP53*), DNA damage response (*RNF4, FANCD2*), and transcription processing (*SIX1, MEPCE, FIP1L1*). Canonical pathway enrichment analysis revealed over-representation of eight pathways (p<0.05) (Supplementary Table 15).

## DISCUSSION

This is the first large-scale GxE analysis for AHI, with the goal of uncovering gene targets for a deeper understanding of the pathophysiology of obstructive sleep apnea. This study examines the effect of EDS on AHI-associated genetic variants in order to elucidate any important biological targets or pathways for the excessively sleepy clinical OSA subtype. We identified significant interactions at 16 genes, of which the following are previously unreported for OSA pathophysiology: *CCDC3, MARCHF1, TMEM26, CPSF4L, MED31, YY1, RAP1GAP*, and *PI4K2B*. This study’s findings suggest the potential of thiamine and resveratrol supplementation for use in OSA patients experiencing excessive daytime sleepiness. Bioinformatics analysis identified cross-trait associations with cardiometabolic traits and medication use, suggesting pathways that may be pertinent to investigate for clinical utility in sleepy OSA patients, given their elevated cardiovascular risk.

Thiamine (vitamin B1) metabolism is highlighted by genes mapped to the *MARCHF1* and *CCDC3* genetic loci from the enrichment of thiamine pyrophosphate and transketolase terms in STRINGdb. Thiamine deficiency can result from high calorie malnutrition, increased age, and gastrointestinal tract factors ^19^. Thiamine deficiency is promoted by fluid loss or antacids use – which can occur in OSA patients due to nighttime sweating or treatment of comorbid gastroesophageal reflux disorder (GERD) ^20^. Thiamine deficiency is also linked to long sleep duration (which is associated with EDS or high sleep propensity), connected to alterations in adenosine triphosphate production^21,22^. Thiamine-containing supplements demonstrate improvement in sleep disturbance and insomnia symptoms ^22^. Thiamine is notably a potent inhibitor of human carbonic anhydrase II with activity comparable to acetazolamide – a medication shown to both lower blood pressure and vascular stiffness, as well as improve AHI and ventilatory instability, in central and obstructive sleep apnea ^23–25^.

*NPY5R* antagonism, resveratrol mechanism, and insulin sensitivity mechanisms may also be important to investigate. *MARCHF1,* the primary mapped gene of the *rs13118183* locus is a regulator of insulin sensitivity, controlling cell surface insulin receptor degradation as a ubiquitin ligase ^26^. This is important as insulin resistance has been identified as an antecedent risk factor for OSA and associated with ventilatory control abnormalities, increased upper airway fat, and increased upper airway collapsibility (an endotype associated with both EDS and inflammation) ^7,27,28^. *MARCHF1* gene expression has been noted to decrease in response to resveratrol, a polyphenol supplement that has anti-inflammatory, antioxidant, and estrogen modulator effects ^29,30^. In the context of OSA, resveratrol has been prior suggested for consideration of clinical use in treating both OSA and cancer ^29,30^. Resveratrol is able to induce *SIRT1* activity (downregulated in OSA), alter insulin sensitivity in visceral white adipose tissue that can occur from sleep fragmentation, and reduce myocardial injury that can occur from chronic intermittent hypoxia ^31–33^. In vitro studies report resveratrol’s inhibitory activity against type II phosphatidylinositol 4-kinases, which supports the previously unreported gene for OSA this study identified for male sex in rare variant gene-set analysis - *PI4K2B* ^34^. In addition to *MARCHF1*, *NPY1R* and *NPY5R* genes mapped to the *rs13118183* locus which revealed through PPI enriched terms a closely connected family: neuropeptide Y (*NPY*) - peptide YY (*PYY*) - and pancreatic polypeptide (*PP*). Notably *NPY* is a vasoconstrictive neuropeptide linked to sleep-wake behavior and may be connected to OSA through its roles in insulin resistance, inflammation, and vascular remodeling ^35,36^. *PYY* is sensitive to sleep duration and energy intake and postulated to be involved in obesity development through circadian disruption ^37^. Drug-gene interaction reported in DGIdb supports this as *NPY5R* is the pharmacological target of velneperit, an investigational obesity drug with anorectic effects.

The strength of this study is in it being the first to assess on a genome-wide scale the interaction role of EDS using two approaches – common variant association analysis, and rare variant gene-based analysis. This work included multi-ethnic source data, sex-specific analyses, and results from multiple bioinformatics tools. One limitation of this work is utilization of race/ethnicity opposed to genetic ancestry for population group definitions. In addition polysomnography based on a single night may result in some misclassification and self-reported EDS can vary over time. Classification of EDS based on self-report data is subjective although the most widely utilized questionnaire for EDS that discriminates sleep disorders groups was used. Unfortunately we were unable to replicate the significant variant and genes identified in WGS discovery results in imputed data. Intra-variability in EDS prevalence was observed within specific ancestries (elevated in HIS, relatively stable in EUR). Thus future ancestry-specific WGS analyses with a large enough sample size for adequate statistical power to enable ancestry-specific analyses would be invaluable. It is difficult to disentangle the role of EDS with respect to OSA – as a symptom, or separate entity. The results here suggest potential gene targets of exploration for the excessively sleepy clinical subtype of OSA – but cannot definitively identify evidence of exact pathways without future validation.

In conclusion, incorporating EDS interactions enabled discovery of genes for OSA that were not revealed by traditional GWAS. This GxE modeling approach assists with precision sleep medicine, as therapeutic designs could differ by exposures or disorder subtypes. Our findings suggest resveratrol and thiamine as supplements for the excessive sleepy subtype of OSA given their modulating pathways. This study’s identified gene targets may help address the complexity inherent to obstructive sleep apnea pathophysiology.

## METHODS

Figure 1 displays the analysis design workflow with cohort description details available in the Supplementary Note. Each contributing study had its protocol approved by the respective Institutional Review Board and participants provided written informed consent.

**Figure 1.**
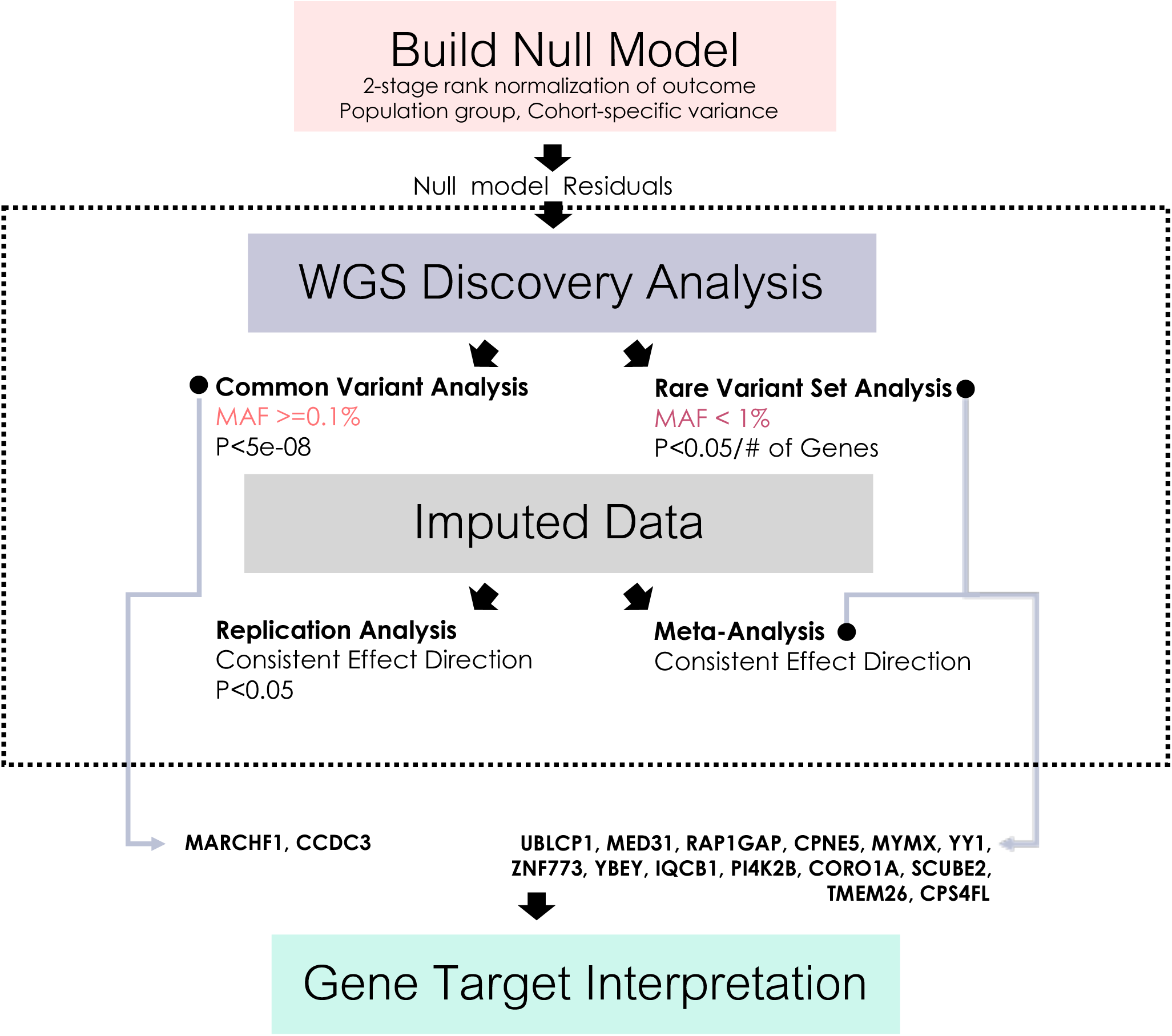
Overall conceptual workflow of this study.

**Figure 2.**
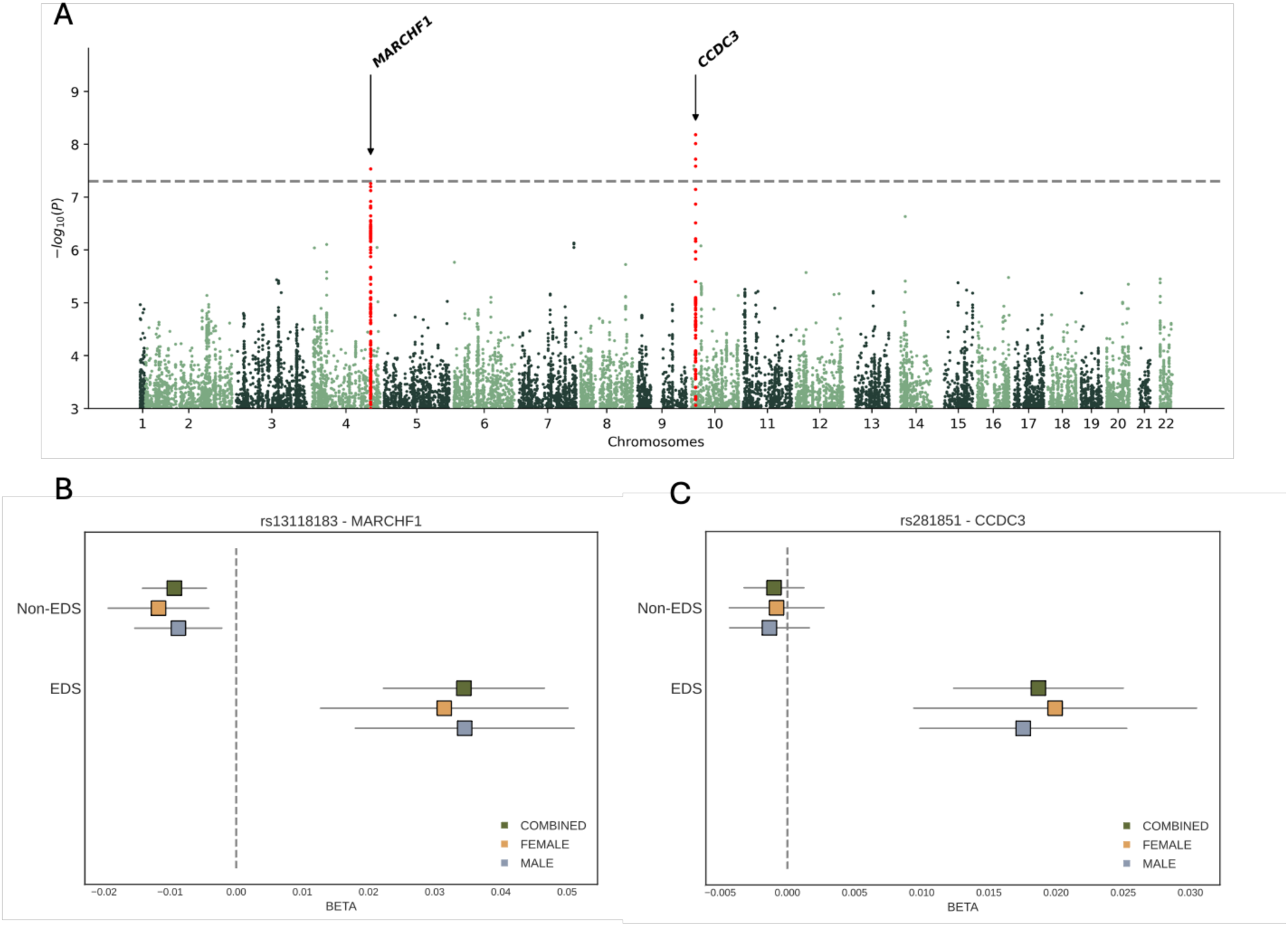
Common variant x EDS interaction effect on AHI. A. Manhattan plot of 1df GxE interaction effect. Two common genetic variants are significant at the genome-wide level (p<5e-08) –rs13118183 (MARCHF1) and rs281851 (CCDC3) – for their interaction effect with EDS. B. Forest plot of rs13118183 (MARCHF1) association with AHI in EDS and non-EDS groups, stratified by sex. C. Forest plot of rs281851 (CCDC3) association with AHI in EDS and non-EDS groups, stratified by sex.

### Data Preparation

For the discovery analysis, whole-genome sequencing (WGS) TOPMed Freeze 8 data from 11,619 individuals (15% AFR, 2% ASN, 28% EUR, 55% HIS) from seven cohorts (Supplementary Table 1) was utilized. Apnea-hypopnea index (AHI) with >=3% oxygen desaturation was retrieved from each cohort. Excessive daytime sleepiness (EDS) was modeled as a binary term from the Epworth Sleepiness Scale (>10: 1, <=10: 0). Age, sex, and body mass index (BMI) were measures obtained at the time of sleep recording. Race/ethnicity measures were derived from dbGaP harmonized demographic data. Genotype data was restricted to minimum sequencing depth of 10, polymorphic PASS only variants, and missingness rate <=5%. A full description of TOPMed whole-genome sequencing can be found at https://topmed.nhlbi.nih.gov/topmed-whole-genome-sequencing-methods-freeze-8. For replication analysis, TOPMed-imputed genotype data was prepared using the TOPMed Imputation Server powered by Minimac4, with retention of variants with imputation quality >=0.4.

### Gene-Environment Interaction Analysis Model

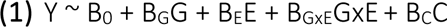

The gene x environment interaction (GxE) model in equation (1) denotes Y as the continuous outcome, apnea-hypopnea index (AHI). The interacting environment term (E) is excessive daytime sleepiness (EDS), defined by the Epworth Sleepiness Scale (>10: 1, <=10: 0). C denotes random effects (PC-Relate estimated kinship matrix, household matrix for HCHS/SOL) and fixed effects (age, sex, BMI, age x EDS, sex x EDS, BMI x EDS, 10 PC-AiR PCs for ancestral group population structure, and race/ethnicity-specific cohort) ^38,39^. With this model design, GENESIS (v2.22.2) was first used to retrieve residuals from the null model allowing for race/ethnicity- and cohort-specific variance, with fully-adjusted two-stage rank normalizawon of AHI with the norm=’ALL’ and rescale=’residSD’ parameters ^40^.

### Common Variant Analysis

For common variant WGS discovery analysis, GEM (v.1.5.2) soxware was used to conduct common variant (minor allele frequency [MAF] ≥0.1%) associawon analysis on the GENESIS output residuals with robust standard error estimation (--robust 1) and no outcome centering (--center 0) ^41^. Summary statistics were retrieved from the following tests: 1 degree-of-freedom (df) genetic effect (B_G_), 1df GxE interaction effect (B_GxE_), and the 2df joint G,GxE effect (B_G_, B_GxE_). Significant variants were those passing genome-wide significance level (p < 5e-8). Summary stawswcs were processed using EasyQC2 to remove variants with missing and out of range values^42^.

### Rare Variant Set-Based Analysis

For rare variant set-based analysis, first variants with MAF<1% were aggregated into genes based on GENCODEv28, restricting variants to those marked as high-confidence non-synonymous loss of function, damaging or deleterious missense (by SIFT4G, Polyphen2 HumDiv, Polyphen2 HumVar, or LRT), or in-frame insertion-deletion with positive FATHMM-XF coding score ^43–46^. Following this, MAGEE (v1.2.0) was used to conduct rare variant gene-based analyses enforcing the double Fisher’s method (tests==‘JD’)^47^. P-value summary statistics output for the main genetic effect (“MF” test), interaction effect (“IF” test), and the joint effect (“JD” test) were retrieved. Summary statistics were processed using EasyQC2 to remove variants with missing and out of range values and inflation corrected ^42^.

### Variant and Gene Prioritization

For common variant analysis, FUMA SNP2GENE (v.1.5.2) was used to filter significant (P<5×10^-8^) signals found on the same chromosome to independent loci defined by lead SNPs using distance criteria (500 kilobases) and linkage disequilibrium (r^2^<0.1) from the 1000G Phase 3 *ALL* panel ^48^. The Open Targets Genetics platform was then used to map each final lead variant to its primary mapped gene by prioritizing firstly direct gene overlap (e.g. intronic), followed by nearest transcription start site, or lastly highest V2G score^49^. For rare variant set-based analysis, significant genes were those with p<3e-6, accounting for the total number of tested genes.

### Replication and Meta-Analysis

After WGS discovery analysis, variants in common variant analysis and genes from rare variant set analysis were checked for replication (P<0.05) by executing the same analyses in 8,904 separate TOPMed-imputed samples from 7 cohorts (Supplementary Table 1) comprising of 3 population groups (1% AFR, 52% EUR, 47% HIS). Common variant meta-analysis was conducted on the WGS and imputed summary statistics using METAL (v2010-02-08), utilizing SCHEME INTERACTION (https://genome.sph.umich.edu/wiki/Meta_Analysis_of_SNPxEnvironment_Interaction) for the 2df joint G,GxE test and SCHEME STDERR for the 1df tests. Rare variant set-based meta-analysis was conducted using MAGEE software ^47,50–52^.

### Final Genes and Variants

The above steps were repeated for pooled sex, female sex, and male sex analysis. The final prioritized genomic loci were (a) significant by the 1df GxE interaction test or (b) significant by the G,GxE joint test with stronger GxE interaction signal (P_G_ > P_GxE_).

### Bioinformatics Analysis

Post-GxE analysis first included annotating whether final genes and variants for OSA have been previously reported, by querying PheWeb (https://pheweb.org/UKB-TOPMed/), GWAS Catalog (https://www.ebi.ac.uk/gwas/), Sleep Disorders Knowledge Portal (https://sleep.hugeamp.org) and four large-scale prior genomic analyses ^3,16–18^. Next, aforementioned FUMA SNP2GENE analysis output was processed to denote the extended gene loci for each lead variant identified in the common variant association analysis, and any enriched tissues defined by MAGMA. FUMA SNP2GENE specifically identifies extended gene loci for each lead variant identified in individual common variant association analysis, based on significant (FDR<0.05) cis-eQTL associations up to 1Mb away and significant (false discovery rate (FDR) <1e-6) chromatin interactions with genes 250bp upstream or 500bp downstream of the transcription start site (TSS). STRINGdb (v12.0) was used to investigate each of these corresponding gene loci by pathway/ontology enrichment analysis from the database’s identified protein-protein interactions. Third, the set of final primary genes from the rare variant and common variant analyses were queried in Open Target Genetics (v22.1) to identify any strong cross-trait associations (Locus-to-Gene score>=0.7, p<5e-08) and medication-related traits (p<5e-08) for therapeutic context; queried in Open Targets Platform (v.24.03) to understand mouse model functional effect; queried in DGIdb (v.5.0) for noting druggable gene targets (interaction score >=1.0); and analyzed by QIAGEN Ingenuity Pathway Analysis ^48,49,53–57^.

## Data Availability

All data produced in the present study are available upon reasonable request to the authors.

## Acknowledgements

This work was supported by the National Institute of Health (NIH) grants R01HL153814 (to H.W.) and R35HL135818 (to S.R.).

Molecular data for the Trans-Omics in Precision Medicine (TOPMed) program was supported by the National Heart, Lung and Blood Institute (NHLBI). Whole-genome sequencing for “NHLBI TOPMed: The Cleveland Family Study (CFS)” (phs000954) was performed at the University of Washington Northwest Genomics Center (3R01HL098433-05S1). Whole genome sequencing for “NHLBI TOPMed - NHGRI CCDG: Atherosclerosis Risk in Communities (ARIC)” (phs001211) was performed at Baylor College of Medicine Human Genome Sequencing Center and Broad Institute of MIT and Harvard (3U54HG003273-12S2/HHSN268201500015C, 3R01HL092577-06S1). Whole-genome sequencing for “NHLBI TOPMed - NHGRI CCDG: Hispanic Community Health Study/Study of Latinos (HCHS/SOL)” (phs001395) was performed at Baylor College of Medicine Human Genome Sequencing Center (HHSN268201600033I). Whole-genome sequencing for “NHLBI TOPMed: Genomic Activities such as Whole Genome Sequencing and Related Phenotypes in the Framingham Heart Study” (phs000974) was performed at Broad Institute of MIT and Harvard (3U54HG003067-12S2). Whole-genome sequencing for “NHLBI TOPMed: NHLBI TOPMed: MESA” (phs001416) was performed at Broad Institute of MIT and Harvard (3U54HG003067-13S1). Whole-genome sequencing for “NHLBI TOPMed: Trans-Omics for Precision Medicine (TOPMed) Whole Genome Sequencing Project: Cardiovascular Health Study” (phs001368) was performed at Baylor College of Medicine Human Genome Sequencing Center (HHSN268201600033I, 3U54HG003273-12S2/HHSN268201500015C). Whole-genome sequencing for “NHLBI TOPMed: The Jackson Heart Study” (phs000964) was performed at University of Washington Northwest Genomics Center (HHSN268201100037C). Core support including centralized genomic read mapping and genotype calling, along with variant quality metrics and filtering were provided by the TOPMed Informatics Research Center (3R01HL-117626-02S1; contract HHSN268201800002I). Core support including phenotype harmonization, data management, sample-identity QC, and general program coordination were provided by the TOPMed Data Coordinating Center (R01HL-120393; U01HL-120393; contract HHSN268201800001I).

The Atherosclerosis Risk in Communities study has been funded in whole or in part with Federal funds from the National Heart, Lung, and Blood Institute, National Institutes of Health, Department of Health and Human Services, under Contract nos. (75N92022D00001, 75N92022D00002, 75N92022D00003, 75N92022D00004, 75N92022D00005). Funding was also supported by R01HL087641 and R01HL086694; National Human Genome Research Institute contract U01HG004402; and National Institutes of Health contract HHSN268200625226C. Infrastructure was partly supported by Grant Number UL1RR025005, a component of the National Institutes of Health and NIH Roadmap for Medical Research. The Genome Sequencing Program (GSP) was funded by the National Human Genome Research Institute (NHGRI), the National Heart, Lung, and Blood Institute (NHLBI), and the National Eye Institute (NEI). The GSP Coordinating Center (U24 HG008956) contributed to cross program scientific initiatives and provided logistical and general study coordination. The Centers for Common Disease Genomics (CCDG) program was supported by NHGRI and NHLBI, and whole genome sequencing was performed at the Baylor College of Medicine Human Genome Sequencing Center (UM1 HG008898). The authors thank the staff and participants of the ARIC study for their important contributions.

The Cleveland Family Study has been supported in part by National Institutes of Health grants [R01-HL046380, KL2-RR024990, R35-HL135818, and R01-HL113338].

Cardiovascular Health Study was supported by NHLBI contracts HHSN268201200036C, HHSN268200800007C, HHSN268201800001C, N01HC55222, N01HC85079, N01HC85080, N01HC85081, N01HC85082, N01HC85083, N01HC85086, 75N92021D00006; and NHLBI grants U01HL080295, R01HL085251, R01HL087652, R01HL105756, R01HL103612, R01HL120393, and U01HL130114 with additional contribution from the National Institute of Neurological Disorders and Stroke (NINDS). Additional support was provided through R01AG023629 from the National Institute on Aging (NIA). A full list of principal CHS investigators and institutions can be found at CHS-NHLBI.org. The content is solely the responsibility of the authors and does not necessarily represent the official views of the National Institutes of Health.

The Framingham Heart Study (FHS) acknowledges the support of contracts NO1-HC-25195, HHSN268201500001I and 75N92019D00031 from the National Heart, Lung and Blood Institute and grant supplement R01 HL092577-06S1 for this research. We also acknowledge the dedication of the FHS study participants without whom this research would not be possible. Dr. Vasan is supported in part by the Evans Medical Foundation and the Jay and Louis Coffman Endowment from the Department of Medicine, Boston University School of Medicine.

The Hispanic Community Health Study/Study of Latinos (HCHS/SOL) is a collaborative study supported by contracts from the National Heart, Lung, and Blood Institute (NHLBI) to the University of North Carolina (HHSN268201300001I / N01-HC-65233), University of Miami (HHSN268201300004I / N01-HC-65234), Albert Einstein College of Medicine (HHSN268201300002I / N01-HC-65235), University of Illinois at Chicago (HHSN268201300003I / N01-HC-65236 Northwestern Univ), and San Diego State University (HHSN268201300005I / N01-HC-65237). The following Institutes/Centers/Offices have contributed to the HCHS/SOL through a transfer of funds to the NHLBI: National Institute on Minority Health and Health Disparities, National Institute on Deafness and Other Communication Disorders, National Institute of Dental and Craniofacial Research, National Institute of Diabetes and Digestive and Kidney Diseases, National Institute of Neurological Disorders and Stroke, NIH Institution-Office of Dietary Supplements. The Genetic Analysis Center at the University of Washington was supported by NHLBI and NIDCR contracts (HHSN268201300005C AM03 and MOD03).

MESA and the MESA SHARe project are conducted and supported by the National Heart, Lung, and Blood Institute (NHLBI) in collaboration with MESA investigators. Support for MESA is provided by contracts HHSN268201500003I, N01-HC-95159, N01-HC-95160, N01-HC-95161, N01-HC-95162, N01-HC-95163, N01-HC-95164, N01-HC-95165, N01-HC-95166, N01-HC-95167, N01-HC-95168, N01-HC-95169, UL1-TR-000040, UL1-TR-001079, UL1-TR-001420. MESA Family is conducted and supported by the National Heart, Lung, and Blood Institute (NHLBI) in collaboration with MESA investigators. Support is provided by grants and contracts R01HL071051, R01HL071205, R01HL071250, R01HL071251, R01HL071258, R01HL071259, and by the National Center for Research Resources, Grant UL1RR033176. The provision of genotyping data was supported in part by the National Center for Advancing Translational Sciences, CTSI grant UL1TR001881, and the National Institute of Diabetes and Digestive and Kidney Disease Diabetes Research Center (DRC) grant DK063491 to the Southern California Diabetes Endocrinology Research Center.

The Osteoporotic Fractures in Men (MrOS) Study is supported by NIH funding. The following institutes provide support: the National Institute on Aging (NIA), the National Institute of Arthritis and Musculoskeletal and Skin Diseases (NIAMS), NCATS, and NIH Roadmap for Medical Research under the following grant numbers: U01 AG027810, U01 AG042124, U01 AG042139, U01 AG042140, U01 AG042143, U01 AG042145, U01 AG042168, U01 AR066160, and UL1 TR000128. The NHLBI provides funding for the MrOS Sleep ancillary study “Outcomes of Sleep Disorders in Older Men” under the following grant numbers: R01 HL071194, R01 HL070848, R01 HL070847, R01 HL070842, R01 HL070841, R01 HL070837, R01 HL070838, and R01 HL070839. The NIAMS provides funding for the MrOS ancillary study ‘Replication of candidate gene associations and bone strength phenotype in MrOS’ under the grant number R01 AR051124. The NIAMS provides funding for the MrOS ancillary study ‘GWAS in MrOS and SOF’ under the grant number RC2 AR058973.

Funding for the Western Australian Sleep Health Study was obtained from the Sir Charles Gairdner and Hollywood Private Hospital Research Foundations, the Western Australian Sleep Disorders Research Institute, and the Centre for Genetic Epidemiology and Biostatistics at the University of Western Australia. Funding for the GWAS genotyping obtained from the Ontario Institute for Cancer Research and a McLaughlin Centre Accelerator Grant from the University of Toronto.

The Jackson Heart Study (JHS) is supported and conducted in collaboration with Jackson State University (HHSN268201800013I), Tougaloo College (HHSN268201800014I), the Mississippi State Department of Health (HHSN268201800015I) and the University of Mississippi Medical Center (HHSN268201800010I, HHSN268201800011I and HHSN268201800012I) contracts from the National Heart, Lung, and Blood Institute (NHLBI) and the National Institute on Minority Health and Health Disparities (NIMHD). The authors also wish to thank the staffs and participants of the JHS. The views expressed in this manuscript are those of the authors and do not necessarily represent the views of the National Heart, Lung, and Blood Institute; the National Institutes of Health; or the U.S. Department of Health and Human Services.

## Author Contributions

PN, SR, and HW designed this study.

PN, NK, JL, SG, YX, YZ, BS, TF, BEC, TS, HW participated in data analysis.

All authors participated in data acquisition including cohort data preparation and harmonization and/or interpretation of discussion of results.

PN, BC, TS, SR, and HW drafted the manuscript.

All authors reviewed and approved the final version of the paper that was submitted to the journal.

## References

1. Redline, S., Azarbarzin, A. & Peker, Y. Obstructive sleep apnoea heterogeneity and cardiovascular disease. Nat Rev Cardiol 20, 560–573 (2023).

2. Szily, M. et al. Genetic influences on the onset of obstructive sleep apnoea and daytime sleepiness: a twin study. Respir Res 20, 125 (2019).

3. Sofer, T. et al. Genome-wide association study of obstructive sleep apnoea in the Million Veteran Program uncovers genetic heterogeneity by sex. EBioMedicine 90, 104536 (2023).

4. Rosenberg, R., Schweitzer, P.K., Steier, J. & Pepin, J.L. Residual excessive daytime sleepiness in patients treated for obstructive sleep apnea: guidance for assessment, diagnosis, and management. Postgrad Med 133, 772–783 (2021).

5. Lal, C., Weaver, T.E., Bae, C.J. & Strohl, K.P. Excessive Daytime Sleepiness in Obstructive Sleep Apnea. Mechanisms and Clinical Management. Ann Am Thorac Soc 18, 757–768 (2021).

6. Zinchuk, A. & Yaggi, H.K. Phenotypic Subtypes of OSA: A Challenge and Opportunity for Precision Medicine. Chest 157, 403–420 (2020).

7. Huang, T. et al. C-reactive Protein and Risk of OSA in Four US Cohorts. Chest 159, 2439–2448 (2021).

8. Liu, Y. et al. Physical activity, sedentary behaviour and incidence of obstructive sleep apnoea in three prospective US cohorts. Eur Respir J 59(2022).

9. Liu, Y., Tabung, F.K., Stampfer, M.J., Redline, S. & Huang, T. Overall diet quality and proinflammatory diet in relation to risk of obstructive sleep apnea in 3 prospective US cohorts. Am J Clin Nutr 116, 1738–1747 (2022).

10. Reid, M. et al. Association between diet quality and sleep apnea in the Multi-Ethnic Study of Atherosclerosis. Sleep 42(2019).

11. Patel, K., Lawson, M. & Cheung, J. Whole-food plant-based diet reduces daytime sleepiness in patients with OSA. Sleep Med 107, 327–329 (2023).

12. Chasens, E.R., Sereika, S.M., Weaver, T.E. & Umlauf, M.G. Daytime sleepiness, exercise, and physical function in older adults. J Sleep Res 16, 60–5 (2007).

13. Bock, J., Covassin, N. & Somers, V. Excessive daytime sleepiness: an emerging marker of cardiovascular risk. Heart 108, 1761–1766 (2022).

14. Taliun, D. et al. Sequencing of 53,831 diverse genomes from the NHLBI TOPMed Program. Nature 590, 290–299 (2021).

15. Nagarajan, P. et al. 0034 Genetic Variants for Obstructive Sleep Apnea Identified after Modeling Interactions with Daytime Sleepiness. Sleep 46, A15–A16 (2023).

16. Cade, B.E. et al. Whole-genome association analyses of sleep-disordered breathing phenotypes in the NHLBI TOPMed program. Genome Med 13, 136 (2021).

17. Strausz, S. et al. Genetic analysis of obstructive sleep apnoea discovers a strong association with cardiometabolic health. Eur Respir J 57(2021).

18. Xu, H. et al. Genome-Wide Association Study of Obstructive Sleep Apnea and Objective Sleep-related Traits Identifies Novel Risk Loci in Han Chinese Individuals. Am J Respir Crit Care Med 206, 1534–1545 (2022).

19. Dhir, S., Tarasenko, M., Napoli, E. & Giulivi, C. Neurological, Psychiatric, and Biochemical Aspects of Thiamine Deficiency in Children and Adults. Front Psychiatry 10, 207 (2019).

20. Harper, R.M., Kumar, R., Ogren, J.A. & Macey, P.M. Sleep-disordered breathing: effects on brain structure and function. Respir Physiol Neurobiol 188, 383–91 (2013).

21. Hernandez-Vazquez, A.J. et al. Thiamine Deprivation Produces a Liver ATP Deficit and Metabolic and Genomic Effects in Mice: Findings Are Parallel to Those of Biotin Deficiency and Have Implications for Energy Disorders. J Nutrigenet Nutrigenomics 9, 287–299 (2016).

22. De Simone, M., De Feo, R., Choucha, A., Ciaglia, E. & Fezeu, F. Enhancing Sleep Quality: Assessing the Efficacy of a Fixed Combination of Linden, Hawthorn, Vitamin B1, and Melatonin. Med Sci (Basel) 12(2023).

23. Ozdemir, Z.O., Senturk, M. & Ekinci, D. Inhibition of mammalian carbonic anhydrase isoforms I, II and VI with thiamine and thiamine-like molecules. J Enzyme Inhib Med Chem 28, 316–9 (2013).

24. Eskandari, D., Zou, D., Grote, L., Hoff, E. & Hedner, J. Acetazolamide Reduces Blood Pressure and Sleep-Disordered Breathing in Patients With Hypertension and Obstructive Sleep Apnea: A Randomized Controlled Trial. J Clin Sleep Med 14, 309–317 (2018).

25. Schmickl, C.N. et al. Acetazolamide for OSA and Central Sleep Apnea: A Comprehensive Systematic Review and Meta-Analysis. Chest 158, 2632–2645 (2020).

26. Nagarajan, A. et al. MARCH1 regulates insulin sensitivity by controlling cell surface insulin receptor levels. Nat Commun 7, 12639 (2016).

27. Huang, T. et al. Insulin Resistance, Hyperglycemia, and Risk of Developing Obstructive Sleep Apnea in Men and Women in the United States. Ann Am Thorac Soc 19, 1740–1749 (2022).

28. Llanos, O.L. et al. Pharyngeal collapsibility during sleep is elevated in insulin-resistant females with morbid obesity. Eur Respir J 47, 1718–26 (2016).

29. Dai, H. et al. Resveratrol inhibits the malignant progression of hepatocellular carcinoma via MARCH1-induced regulation of PTEN/AKT signaling. Aging (Albany NY) 12, 11717–11731 (2020).

30. Porcacchia, A.S., Moreira, G.A., Andersen, M.L. & Tufik, S. The use of resveratrol in the treatment of obstructive sleep apnea and cancer: a commentary on common targets. J Clin Sleep Med 18, 333–334 (2022).

31. Carreras, A. et al. Effect of resveratrol on visceral white adipose tissue inflammation and insulin sensitivity in a mouse model of sleep apnea. Int J Obes (Lond) 39, 418–23 (2015).

32. Sun, Z.M. et al. Resveratrol protects against CIH-induced myocardial injury by targeting Nrf2 and blocking NLRP3 inflammasome activation. Life Sci 245, 117362 (2020).

33. Malicki, M., Karuga, F.F., Szmyd, B., Sochal, M. & Gabryelska, A. Obstructive Sleep Apnea, Circadian Clock Disruption, and Metabolic Consequences. Metabolites 13(2022).

34. Srivastava, R. et al. Resveratrol inhibits type II phosphatidylinositol 4-kinase: a key component in pathways of phosphoinositide turn over. Biochem Pharmacol 70, 1048–55 (2005).

35. Li, M.M., Zheng, Y.L., Wang, W.D., Lin, S. & Lin, H.L. Neuropeptide Y: An Update on the Mechanism Underlying Chronic Intermittent Hypoxia-Induced Endothelial Dysfunction. Front Physiol 12, 712281 (2021).

36. Shen, Y.C. et al. Roles of Neuropeptides in Sleep-Wake Regulation. Int J Mol Sci 23(2022).

37. Chaput, J.P. et al. The role of insufficient sleep and circadian misalignment in obesity. Nat Rev Endocrinol 19, 82–97 (2023).

38. Conomos, M.P., Reiner, A.P., Weir, B.S. & Thornton, T.A. Model-free Estimation of Recent Genetic Relatedness. Am J Hum Genet 98, 127–48 (2016).

39. Conomos, M.P., Miller, M.B. & Thornton, T.A. Robust inference of population structure for ancestry prediction and correction of stratification in the presence of relatedness. Genet Epidemiol 39, 276–93 (2015).

40. Sofer, T. et al. A fully adjusted two-stage procedure for rank-normalization in genetic association studies. Genet Epidemiol 43, 263–275 (2019).

41. Westerman, K.E., et al. GEM: scalable and flexible gene-environment interaction analysis in millions of samples. Bioinformatics 37, 3514-3520 (2021).

42. Winkler, T.W. et al. Quality control and conduct of genome-wide association meta-analyses. Nat Protoc 9, 1192–212 (2014).

43. Vaser, R., Adusumalli, S., Leng, S.N., Sikic, M. & Ng, P.C. SIFT missense predictions for genomes. Nat Protoc 11, 1–9 (2016).

44. Adzhubei, I., Jordan, D.M. & Sunyaev, S.R. Predicting functional effect of human missense mutations using PolyPhen-2. Curr Protoc Hum Genet Chapter 7, Unit7 20 (2013).

45. Chun, S. & Fay, J.C. Identification of deleterious mutations within three human genomes. Genome Res 19, 1553–61 (2009).

46. Rogers, M.F. et al. FATHMM-XF: accurate prediction of pathogenic point mutations via extended features. Bioinformatics 34, 511–513 (2018).

47. Wang, X. et al. Efficient gene-environment interaction tests for large biobank-scale sequencing studies. Genet Epidemiol 44, 908–923 (2020).

48. Watanabe, K., Taskesen, E., van Bochoven, A. & Posthuma, D. Functional mapping and annotation of genetic associations with FUMA. Nat Commun 8, 1826 (2017).

49. Ghoussaini, M. et al. Open Targets Genetics: systematic identification of trait-associated genes using large-scale genetics and functional genomics. Nucleic Acids Res 49, D1311–D1320 (2021).

50. Willer, C.J., Li, Y. & Abecasis, G.R. METAL: fast and efficient meta-analysis of genomewide association scans. Bioinformatics 26, 2190–1 (2010).

51. Manning, A.K. et al. Meta-analysis of gene-environment interaction: joint estimation of SNP and SNP x environment regression coefficients. Genet Epidemiol 35, 11–8 (2011).

52. Wang, X. et al. Genomic summary statistics and meta-analysis for set-based gene-environment interaction tests in large-scale sequencing studies. (medRxiv, 2022).

53. de Leeuw, C.A., Mooij, J.M., Heskes, T. & Posthuma, D. MAGMA: generalized gene-set analysis of GWAS data. PLoS Comput Biol 11, e1004219 (2015).

54. Szklarczyk, D. et al. STRING v11: protein-protein association networks with increased coverage, supporting functional discovery in genome-wide experimental datasets. Nucleic Acids Res 47, D607–D613 (2019).

55. Freshour, S.L. et al. Integration of the Drug-Gene Interaction Database (DGIdb 4.0) with open crowdsource efforts. Nucleic Acids Res 49, D1144–D1151 (2021).

56. Koscielny, G. et al. Open Targets: a platform for therapeutic target identification and validation. Nucleic Acids Res 45, D985–D994 (2017).

57. Kramer, A., Green, J., Pollard, J., Jr. & Tugendreich, S. Causal analysis approaches in Ingenuity Pathway Analysis. Bioinformatics 30, 523–30 (2014).

