## Supplementary Tables for "Gene-Excessive Sleepiness Interactions Suggest Treatment Targets for Obstructive Sleep Apnea Subtype"

**Supplementary Table 1. Data Characteristics**

| *Cohort* | Population Group | N | EDS Presence | Females | Age | BMI | AHI |
|---|---|---|---|---|---|---|---|
| | | | N (%) | N (%) | Mean (SD) | Mean, (SD) | Mean (SD) |
| **Whole Genome Sequencing Data** | | | | | | | |
| *ARIC* | EUR | 1083 | 311 (14.4%) | 575 (8.77%) | 62.19 (5.70) | 28.79 (5.07) | 13.65 (14.20) |
| *HCHS/SOL* | HIS | 5927 | 902 (41.7%) | 3426 (52.27%) | 46.73 (13.87) | 30.06 (6.36) | 6.76 (12.72) |
| *CFS* | EUR | 481 | 138 (6.4%) | 242 (3.69%) | 43.60 (19.69) | 30.86 (8.87) | 17.96 (24.76) |
| | AFR | 505 | 184 (8.5%) | 285 (4.35%) | 38.96 (19.08) | 32.38 (9.46) | 19.11 (26.80) |
| *FHS* | EUR | 464 | 86 (4.0%) | 231 (3.52%) | 60.01 (8.50) | 28.49 (5.13) | 12.36 (13.31) |
| *MESA* | AFR | 488 | 88 (4.1%) | 263 (4.01%) | 68.67 (9.08) | 30.28 (5.68) | 19.48 (19.70) |
| | EUR | 694 | 77 (3.6%) | 370 (5.65%) | 68.53 (9.08) | 27.89 (5.09) | 18.39 (17.80) |
| | HIS | 454 | 57 (2.6%) | 240 (3.66%) | 68.29 (9.21) | 30.08 (5.46) | 21.73 (18.51) |
| | EAS | 223 | 27 (1.2%) | 113 (1.72%) | 67.74 (9.06) | 24.25 (3.29) | 20.54 (18.70) |
| *CHS* | EUR | 540 | 109 (5.0%) | 318 (4.85%) | 77.72 (4.24) | 27.30 (4.43) | 15.36 (15.61) |
| | AFR | 193 | 59 (2.7%) | 120 (1.83%) | 75.69 (4.67) | 28.64 (4.93) | 15.26 (15.02) |
| *JHS* | AFR | 567 | 126 (5.8%) | 371 (5.66%) | 63.45 (10.88) | 31.87 (6.89) | 15.69 (15.93) |
| *TOTAL* | | **11619** | **2164** | **6554** | | | |
| **Imputed Genotype Data** | | | | | | | |
| *ARIC* | EUR | 457 | 133 (7.30%) | 229 (6.28%) | 62.90 (5.68) | 28.97 (5.14) | 14.76 (16.84) |
| *CFS* | AFR | 72 | 16 (0.88%) | 39 (1.07%) | 26.19 (18.18) | 26.79 (9.61) | 7.96 (15.28) |
| | EUR | 103 | 19 (1.04%) | 55 (1.51%) | 38.11 (17.83) | 29.47 (7.13) | 13.91 (21.06) |
| *CHS* | EUR | 216 | 51 (2.80%) | 140 (3.84%) | 77.74 (4.41) | 26.84 (4.40) | 15.08 (14.00) |
| | AFR | 8 | 4 (0.22%) | 1 (0.03%) | 75.50 (5.37) | 28.10 (2.89) | 23.16 (14.23) |
| *FHS* | EUR | 139 | 27 (1.48%) | 69 (1.89%) | 56.55 (9.74) | 28.54 (4.75) | 13.90 (16.19) |
| *WASHS* | EUR | 1493 | 648 (35.5%) | 607 (16.66%) | 52.30 (13.77) | 31.86 (7.89) | 14.43 (18.68) |
| *MrOS* | EUR | 2209 | 279 (15.3%) | 0 (0.00%) | 76.68 (5.67) | 27.22 (3.74) | 17.31 (15.49) |
| *HCHS/SOL* | HIS | 4207 | 646 (35.4%) | 2504 (68.72%) | 45.32 (13.69) | 29.33 (5.35) | 5.91 (11.37) |
| *TOTAL* | | **8904** | **1823** | **3644** | | | |

**Supplementary Table 2. Extended Gene Loci Identified by FUMA SNP2GENE – Interaction Effect, WGS Discovery Analysis**

| Gene | eQTL | eQTL Association Tissues | Position | Chromatin Interaction | Chromatin Interaction Tissues | Lead Variant |
|---|---|---|---|---|---|---|
| ***NAF1*** | 0 | | 0 | Yes | Mesenchymal_Stem_Cell | rs13118183 |
| ***NPY1R*** | 0 | | 0 | Yes | Mesenchymal_Stem_Cell | rs13118183 |
| ***NPY5R*** | 1 | CMC_SVA_cis | 0 | Yes | Mesenchymal_Stem_Cell | rs13118183 |
| ***TKTL2*** | 0 | | 0 | Yes | Mesendoderm:hESC | rs13118183 |
| ***MARCHF1*** | 21 | CMC_NoSVA_cis, GTEx/v8/Adipose_Visceral_Omentum | 218 | Yes | Dorsolateral_Prefrontal_Cortex, Left_Ventricle, Liver, IMR90, Mesenchymal_Stem_Cell, Mesendoderm, Neural_Progenitor_Cell, Trophoblast-like_Cell:hESC | rs13118183 |
| ***FAM218A*** | 0 | | 0 | Yes | Mesendoderm, hESC | rs13118183 |
| ***TRIM60*** | 0 | | 0 | Yes | Promoter_anchored_loops | rs13118183 |
| ***UPF2*** | 0 | | 0 | Yes | Mesenchymal_Stem_Cell | rs281851 |
| ***DHTKD1*** | 0 | | 0 | Yes | Mesenchymal_Stem_Cell | rs281851 |
| ***SEC61A2*** | 0 | | 0 | Yes | Mesenchymal_Stem_Cell | rs281851 |
| ***NUDT5*** | 0 | | 0 | Yes | Left_Ventricle, Liver, GM12878, IMR90, Mesenchymal_Stem_Cell, Mesendoderm, Trophoblast-like_Cell, hESC | rs281851 |
| ***CDC123*** | 0 | | 0 | Yes | Left_Ventricle, Liver, GM12878, IMR90, Mesenchymal_Stem_Cell, Mesendoderm, Trophoblast-like Cell, hESC | rs281851 |
| ***CAMK1D*** | 0 | | 0 | Yes | IMR90, Mesenchymal_Stem_Cell, Mesendoderm, Trophoblast-like_Cell, hESC | rs281851 |
| ***CCDC3*** | 65 | PsychENCODE_eQTLs, CMC_SVA_cis, GTEx/v8/Adipose_Subcutaneous, GTEx/v8/Adipose_Visceral_Omentum, GTEx/v8/Artery_Aorta, GTEx/v8/Artery_Tibial, GTEx/v8/Colon_Sigmoid, GTEx/v8/Heart_Left_Ventricle, GTEx/v8/Lung, GTEx/v8/Nerve_Tibial, GTEx/v8/Cells_Cultured_fibroblasts | 77 | Yes | IMR90, Mesenchymal_Stem_Cell, Mesendoderm, hESC | rs281851 |
| ***OPTN*** | 0 | 2 | 0 | Yes | IMR90, Mesenchymal_Stem_Cell, Mesendoderm, hESC | rs281851 |
| ***PHYH*** | 1 | eQTLGen_cis_eQTLs | 0 | No | | rs281851 |

**Supplementary Table 3. Variation in Number of Variants Mapped to Gene Sets – Rare Variant Set Analysis**

| Gene | *Sex* | *# Variants in WGS* | *# Variants in Imputed* |
|---|---|---|---|
| ***SCUBE2*** | Combined | 25 | 3 |
| ***TMEM26*** | Male | 2 | 0 |
| ***CPSF4L*** | Male | 5 | 1 |

| **Supplementary Table 4. Common Variant Analysis WGS Additional Significant Loci – Main Genetic Effect** | | | | | | | | | | | |
|---|---|---|---|---|---|---|---|---|---|---|---|
| | | | | | | | **MAIN GENETIC EFFECT** | | **INTERACTION EFFECT** | | **JOINT EFFECT** |
| ***rsID*** | **Sex** | **Chr:Pos (b38)** | **Eff/Alt** | **EAF** | **Gene Locus (FUMA SNP2GENE)** | **N** | **Beta (SE)** | **P-Value** | **Beta (SE)** | **P-Value** | **P-Value** |
| *rs35370454** | Combined | 7:112425915 | C/T | 0.99 | *IMMP2L, LRRN3, DOCK4, ZNF277,* ***IFRD1,*** *LSMEM1, TMEM168, C7orf60, GPR85, LINC00998, TSRM* | 11615 | 0.024 (0.004) | **1.32E-08** | -0.028 (0.012) | 2.35E-02 | 9.18E-08 |
| *rs13433190** | Female | 20:3751618 | G/T | 0.93 | *TMEM239, DDRGK1, GFRA4, ADAM33, SIGLEC1,* ***HSPA12B,*** *C20orf27, SPEF1, CENPB, CDC25B, MAVS, RP11-352D3.2* | 6546 | 0.018 (0.003) | **6.07E-09** | -0.012 (0.009) | 1.75E-01 | 3.48E-08 |

* rs35370454 and rs13433190 were not present in imputed genotype data.

**Supplementary Table 5. Rare Variant WGS Set-Based Analysis Additional Significant Results – Main Genetic Effect**

| | | | | | *MAIN GENETIC EFFECT* | *INTERACTION EFFECT* | *JOINT EFFECT* |
|---|---|---|---|---|---|---|---|
| **Gene** | **Chr: Position (b38)** | **Sex** | **Dataset** | **# of Variants** | **P-Value** | **P-Value** | **P-Value** |
| ***HBB**** | 11:5,225,464-5,229,395 | Female | WGS | 1 | **2.75E-06** | 3.27E-02 | 9.89E-06 |
| ***KRR1**** | 12:75,490,863-75,511,636 | Female | WGS | 2 | **3.06E-06** | 6.80E-01 | 2.74E-04 |
| ***PKD2L2*** | 5:137,887,968-137,942,747 | Male | WGS | 9 | 1.19E-05 | 3.77E-03 | **1.42E-06** |
| | | | Imputed | 2 | 7.14E-01 | 8.24E-01 | 9.00E-01 |
| ***UBOX5*** | 20:3,107,573-3,160,196 | Male | WGS | 2 | **9.70E-09** | 1.83E-02 | **2.07E-08** |
| | | | Imputed | 1 | 7.82E-01 | 7.58E-01 | 9.03E-01 |

*Variant sets mapped to HBB and KRR1 were not present in imputed genotype data

**Supplementary Table 6. Rare Variant Set-Based Meta-Analysis Additional Significant Loci – Main Genetic Effect**

| | | | | MAIN GENETIC EFFECT | INTERACTION EFFECT | JOINT EFFECT |
|---|---|---|---|---|---|---|
| **Gene** | **Chr:Pos (b38)** | **Sex** | **Number of Variants** | **P-Value** | **P-Value** | **P-Value** |
| ***UQCC3*** | 11:62,670,273-62,673,686 | Combined | 3 | **3.51E-07** | 1.39E-02 | **1.89E-06** |
| ***C6orf52*** | 6:10,671,418-10,694,797 | Female | 2 | 2.00E-05 | 4.41E-04 | **3.28E-07** |
| ***TCF4*** | 18:55,222,185-55,664,787 | Female | 2 | **4.23E-07** | 3.78E-02 | 2.89E-06 |
| ***ZNF383*** | 19:37,217,926-37,248,740 | Female | 2 | 2.29E-04 | 2.59E-04 | **1.06E-06** |
| ***SYS1*** | 20:45,361,937-45,376,798 | Female | 2 | **1.27E-06** | 1.01E-02 | **1.49E-06** |
| ***OSER1*** | 20:44,195,939-44,210,771 | Female | 2 | **1.46E-06** | 2.94E-01 | 6.40E-05 |
| ***PAPOLG*** | 2:60,756,253-60,802,086 | Male | 3 | **1.70E-06** | 8.00E-01 | 1.03E-04 |
| ***TBX20*** | 7:35,202,430-35,254,100 | Male | 2 | 3.27E-05 | 2.60E-04 | **2.53E-07** |
| ***ACAT1*** | 11:108,116,695-108,147,603 | Male | 1 | **9.84E-07** | 1.31E-04 | **7.47E-09** |
| ***TGDS*** | 13:94,574,054-94,596,242 | Male | 2 | **4.72E-09** | 3.33E-02 | **3.97E-08** |
| ***CHAMP1*** | 13:114,314,482-114,337,626 | Male | 3 | **5.98E-09** | 1.87E-02 | **2.60E-08** |
| ***RPS15A*** | 16:18,781,295-18,790,383 | Male | 2 | **7.75E-09** | 2.59E-02 | **4.52E-08** |
| ***ZNF112*** | 19:44,326,555-44,367,217 | Male | 6 | **8.76E-07** | 8.93E-01 | 6.82E-05 |
| ***PIM3*** | 22:49,960,768-49,964,072 | Male | 2 | **7.62E-09** | 4.68E-02 | **8.31E-08** |

**Supplementary Table 7. Assessing Whether Genomic Loci for OSA Are Previously Unreported**

| Variant or Gene | Unreported | GWAS Catalog | PheWeb | Prior OSA GWAS Studies ** |
|---|---|---|---|---|
| ***rs13118183*** | * | | - | - |
| ***rs281851*** | * | | - | - |
| ***SCUBE2*** | | Total protein levels x insomnia interaction; Personality traits or cognitive traits (multivariate analysis); Waist-hip ratio | - | Min SpO2 [S4]; |
| ***TMEM26*** | * | Total PHF-tau (SNP x SNP interaction); Cognitive ability, years of educational attainment or schizophrenia (pleiotropy); Diffuse plaques (SNP x SNP interaction); Diastolic BP; Systolic BP; PR Interval ; BMI; Adult body size; Body mass index (MTAG) | Hypertension | - |
| ***CPSF4L*** | * | Red cell distribution width | - | - |
| ***UBLCP1*** | | | - | AHI 3% [S4]; Avg Desaturation 0% [S4, S5]; |
| ***MED31*** | * | Systolic BP; Waist circumference adjusted for body mass index | - | - |
| ***RAP1GAP*** | * | Immature fraction of reticulocytes; High light scatter reticulocyte percentage of red cells; High light scatter reticulocyte count; Mean reticulocyte volume; Abdominal aortic calcification levels; | - | - |
| ***CPNE5*** | | Heart rate variability traits (SDNN); Heart rate variability traits (RMSSD); Heart rate variability (standard deviation of normal-to-normal intervals); Heart rate variability (corrected standard deviation of normal-to-normal intervals); Heart rate variability (corrected root mean square of successive differences); Heart rate variability (root mean square of successive differences); Resting heart rate; Heart rate variability traits (pvRSA/HF); Pulse Pressure; Heart rate response to exercise | - | Min SpO2 [S4]; |

| | | | | |
|---|---|---|---|---|
| ***MYMX*** | | Mean spheric corpuscular volume; Diastolic BP; Mean reticulocyte volume | - | Avg SpO2 [S4]; Per90 [S4, S5] |
| ***YY1*** | * | Caudate nucleus volume; Dementia; Pulse Pressure; | Atherosclerosis of aorta | - |
| ***ZNF773*** | | | - | Min SpO2 [S4]; |
| ***YBEY*** | | | - | Avg Desaturation 0% [S4] |
| ***IQCB1*** | | Multiple sclerosis; Mean corpuscular volume; Hemoglobin levels | - | Avg SpO2 [S4, S5]; Min SpO2 [S4, S5]; Per90 [S4, S5]; |
| ***PI4K2B*** | * | | - | - |
| ***CORO1A*** | | Cortical thickness; Brain morphology (MOSTest); Multiple sclerosis; BMI; Whole body fat mass (UKB data field 23100) | - | AHI 3% [S4]; |

**Four prior large-scale OSA-related GWAS studies were assessed: Sofer et al 2023 [PMID: 36989840; Table 1, Supplementary Tables S2-S3,S6, S7-S8 (FDR-BH<=0.05), S9-S17], Cade et al 2021 [PMID: 34446064; Tables 3-5, Supplementary Tables S4-S5 [P<0.01]], Strausz et al 2021 [PMID: 33243845; Table 2, Figures S5,S7], and Xu et al 2022 [PMID: 35819321; Table 1, Table 2, Supplementary Tables E3-E5]. Amongst these studies overlapped genes was identified

| Supplementary Table 8. Stratified Analysis According to Exposure Status – Common Variants – WGS Data | | | | | | | | | | |
|---|---|---|---|---|---|---|---|---|---|---|
| **rsID** | **Exposure Status** | **Sex** | **Chr:Position (b38)** | **Effect Allele / Alternative Allele** | **Effect Allele Frequency** | **Gene** | **N** | **Beta (SE)** | **P** | **Direction of Effect** |
| ***rs13118183*** | EDS | Combined | 4:164200775 | A/G | 0.937 | *MARCHF1* | 2163 | 0.025 (0.006) | 3.30E-05 | + |
| ***rs13118183*** | EDS | Female | 4:164200775 | A/G | 0.933 | *MARCHF1* | 1137 | 0.020 (0.010) | 4.60E-02 | + |
| ***rs13118183*** | EDS | Male | 4:164200775 | A/G | 0.942 | *MARCHF1* | 1026 | 0.026 (0.008) | 6.00E-04 | + |
| ***rs13118183*** | No EDS | Combined | 4:164200775 | A/G | 0.939 | *MARCHF1* | 9451 | -0.009 (0.002) | 5.40E-05 | - |
| ***rs13118183*** | No EDS | Female | 4:164200775 | A/G | 0.939 | *MARCHF1* | 5413 | -0.012 (0.004) | 1.40E-03 | - |
| ***rs13118183*** | No EDS | Male | 4:164200775 | A/G | 0.939 | *MARCHF1* | 4038 | -0.009 (0.003) | 4.30E-03 | - |
| ***rs281851*** | EDS | Combined | 10:12924495 | A/C | 0.605 | *CCDC3* | 2163 | 0.018 (0.003) | **4.46E-09** | + |
| ***rs281851*** | EDS | Female | 10:12924495 | A/C | 0.608 | *CCDC3* | 1137 | 0.019 (0.005) | 1.94E-04 | + |
| ***rs281851*** | EDS | Male | 10:12924495 | A/C | 0.602 | *CCDC3* | 1026 | 0.016 (0.004) | 6.00E-06 | + |
| ***rs281851*** | No EDS | Combined | 10:12924495 | A/C | 0.588 | *CCDC3* | 9452 | -0.001 (0.001) | 3.73E-01 | - |
| ***rs281851*** | No EDS | Female | 10:12924495 | A/C | 0.59 | *CCDC3* | 5414 | -0.001 (0.002) | 6.54E-01 | - |
| ***rs281851*** | No EDS | Male | 10:12924495 | A/C | 0.59 | *CCDC3* | 4038 | -0.001 (0.001) | 3.60E-01 | - |

**Supplementary Table 9. Stratified Meta-Analysis According to Exposure Status – Rare Variants**

| *Gene* | Exposure Status | Sex | Number of Variants | P-Value |
|---|---|---|---|---|
| ***UBLCP1*** | EDS | Combined | 2 | 3.39E-02 |
| ***UBLCP1*** | No EDS | Combined | 15 | 6.53E-01 |
| ***SCUBE2*** | EDS | Combined | 50 | 9.60E-01 |
| ***SCUBE2*** | No EDS | Combined | 99 | 5.90E-01 |
| ***MED31*** | EDS | Combined | 1 | 5.56E-01 |
| ***MED31*** | No EDS | Combined | 6 | 8.09E-01 |
| ***RAP1GAP*** | EDS | Female | 2 | 8.62E-01 |
| ***RAP1GAP*** | No EDS | Female | 2 | 6.43E-01 |
| ***CPNE5*** | EDS | Female | 11 | 3.11E-02 |
| ***CPNE5*** | No EDS | Female | 33 | 7.58E-01 |
| ***MYMX*** | EDS | Female | 2 | 2.02E-02 |
| ***MYMX*** | No EDS | Female | 2 | 3.26E-02 |
| ***YY1*** | EDS | Female | * | * |
| ***YY1*** | No EDS | Female | 2 | 9.63E-01 |
| ***ZNF773*** | EDS | Female | 8 | 2.26E-01 |
| ***ZNF773*** | No EDS | Female | 30 | 3.17E-01 |
| ***YBEY*** | EDS | Female | 3 | 6.99E-01 |
| ***YBEY*** | No EDS | Female | 8 | 7.35E-01 |
| ***IQCB1*** | EDS | Male | 10 | 7.59E-01 |
| ***IQCB1*** | No EDS | Male | 32 | 4.72E-01 |
| ***PI4K2B*** | EDS | Male | 9 | 4.05E-01 |
| ***PI4K2B*** | No EDS | Male | 15 | 5.36E-01 |
| ***TMEM26*** | EDS | Male | 5 | 1.06E-02 |
| ***TMEM26*** | No EDS | Male | 14 | 2.44E-01 |
| ***CORO1A*** | EDS | Male | 4 | 4.70E-01 |
| ***CORO1A*** | No EDS | Male | 13 | 1.98E-01 |
| ***CPSF4L*** | EDS | Male | 9 | 4.96E-01 |
| ***CPSF4L*** | No EDS | Male | 14 | 2.45E-01 |

* Denotes this variant set was not available in this subset of samples.

**Supplementary Table 10. MAGMA Tissue-Enrichment Analysis Significant Results**

| *Sex* | Tissue | Beta | Standard Error | P-Value |
|---|---|---|---|---|
| **Female** | Breast Mammary Tissue | 0.069494 | 0.01739 | **3.24E-05** |
| **Female** | Tibial Nerve | 0.049348 | 0.013949 | **2.03E-04** |

*Bonferroni Threshold: p<9.3e-04

**Supplementary Table 11. MAGMA Gene-Level Analysis Significant Results***

| *Gene* | Position | Sex | Number of Variants | N | Z-Statistic | P-Value |
|---|---|---|---|---|---|---|
| ***NOP53*** | 19:47,745,546-47,757,058 | Combined | 3 | 11615 | 4.7344 | **1.10E-06** |
| ***EYA2*** | 20:46,894,624-47,188,844 | Combined | 18 | 11614 | 4.6463 | **1.69E-06** |
| ***ZNF563*** | 19:12,317,477-12,333,720 | Combined | 4 | 11614 | 4.5472 | **2.72E-06** |
| ***WDR19*** | 4:39,182,504-39,285,810 | Female | 9 | 6551 | 4.6988 | **1.31E-06** |

*Bonferroni Threshold: $p<4.3e-06$

| Supplementary Table 12. Open Targets Platform Identified Mouse Phenotypes | | |
|---|---|---|
| **mouseGene** | **phenotype** | **category** |
| *Marchf1* | decreased interleukin-12 secretion | immune system phenotype |
| *Marchf1* | abnormal B cell physiology | hematopoietic system phenotype,immune system phenotype |
| *Marchf1* | decreased tumor necrosis factor secretion | immune system phenotype |
| *Marchf1* | decreased regulatory T cell number | hematopoietic system phenotype,immune system phenotype |
| *Marchf1* | abnormal T cell differentiation | hematopoietic system phenotype,immune system phenotype |
| *Marchf1* | abnormal dendritic cell antigen presentation | immune system phenotype |
| *Marchf1* | abnormal dendritic cell morphology | hematopoietic system phenotype,immune system phenotype |
| *Marchf1* | abnormal level of surface class II molecules | immune system phenotype |
| *Ccdc3* | increased circulating glucose level | homeostasis/metabolism phenotype |
| *Ccdc3* | abnormal fat pad morphology | adipose tissue phenotype |
| *Ccdc3* | weight loss | growth/size/body region phenotype |
| *Ccdc3* | decreased liver weight | liver/biliary system phenotype |
| *Ccdc3* | increased insulin sensitivity | homeostasis/metabolism phenotype |
| *Ccdc3* | decreased susceptibility to age-related hepatic steatosis | liver/biliary system phenotype |
| *Ccdc3* | abnormal glucose homeostasis | homeostasis/metabolism phenotype |
| *Ccdc3* | decreased white fat cell size | adipose tissue phenotype |
| *Ccdc3* | decreased liver triglyceride level | homeostasis/metabolism phenotype,liver/biliary system phenotype |
| *Ccdc3* | decreased circulating leptin level | homeostasis/metabolism phenotype |
| *Ccdc3* | decreased subcutaneous adipose tissue amount | integument phenotype,adipose tissue phenotype |
| *Ublcp1* | tremors | behavior/neurological phenotype |
| *Med31* | decreased fibroblast proliferation | cellular phenotype |
| *Med31* | abnormal cartilage development | skeleton phenotype |
| *Med31* | pallor | integument phenotype |
| *Med31* | edema | homeostasis/metabolism phenotype |
| *Med31* | embryonic growth retardation | embryo phenotype,growth/size/body region phenotype |
| *Med31* | lethality throughout fetal growth and development, complete penetrance | mortality/aging |
| *Med31* | decreased embryo size | embryo phenotype,growth/size/body region phenotype |
| *Med31* | abnormal embryonic growth/weight/body size | growth/size/body region phenotype |
| *Med31* | open neural tube | embryo phenotype,nervous system phenotype |
| *Med31* | decreased fetal size | growth/size/body region phenotype |
| *Med31* | small thoracic cage | skeleton phenotype |
| *Med31* | spina bifida | embryo phenotype,nervous system phenotype |

| | | |
|---|---|---|
| *Med31* | abnormal skeleton morphology | skeleton phenotype |
| *Med31* | decreased cell proliferation | cellular phenotype |
| *Med31* | abnormal limb development | limbs/digits/tail phenotype |
| *Med31* | small cranium | craniofacial phenotype,skeleton phenotype |
| *Med31* | decreased bone ossification | skeleton phenotype |
| *Scube2* | decreased bone trabecula number | skeleton phenotype |
| *Scube2* | abnormal skeleton development | skeleton phenotype |
| *Scube2* | abnormal bone ossification | skeleton phenotype |
| *Scube2* | decreased trabecular bone volume | skeleton phenotype |
| *Scube2* | abnormal trabecular bone morphology | skeleton phenotype |
| *Scube2* | decreased volumetric bone mineral density | skeleton phenotype |
| *Scube2* | short femur | limbs/digits/tail phenotype,skeleton phenotype |
| *Scube2* | abnormal long bone epiphyseal plate proliferative zone | skeleton phenotype |
| *Scube2* | abnormal endochondral bone ossification | skeleton phenotype |
| *Scube2* | decreased osteoblast cell number | skeleton phenotype |
| *Scube2* | decreased compact bone thickness | skeleton phenotype |
| *Scube2* | decreased chondrocyte proliferation | cellular phenotype,skeleton phenotype |
| *Scube2* | decreased body size | growth/size/body region phenotype |
| *Scube2* | decreased body weight | growth/size/body region phenotype |
| *Scube2* | abnormal chondrocyte differentiation | skeleton phenotype |
| *Scube2* | decreased trabecular bone thickness | skeleton phenotype |
| *Scube2* | abnormal long bone hypertrophic chondrocyte zone | skeleton phenotype |
| *Scube2* | decreased body height | growth/size/body region phenotype |
| *Scube2* | short tibia | limbs/digits/tail phenotype,skeleton phenotype |
| *Scube2* | decreased body length | growth/size/body region phenotype |
| *Eya2* | increased or absent threshold for auditory brainstem response | hearing/vestibular/ear phenotype |
| *Eya2* | enlarged spleen | hematopoietic system phenotype,immune system phenotype,growth/size/body region phenotype |
| *Eya2* | enlarged lymph nodes | immune system phenotype |
| *Eya2* | abnormal spleen morphology | hematopoietic system phenotype,immune system phenotype |
| *Eya2* | abnormal lymph node morphology | immune system phenotype |
| *Eya2* | small thymus | hematopoietic system phenotype,immune system phenotype,endocrine/exocrine gland phenotype |
| *Eya2* | impaired hearing | hearing/vestibular/ear phenotype |
| *Eya2* | abnormal pinna reflex | behavior/neurological phenotype |
| *Eya2* | small spleen | hematopoietic system phenotype,immune system phenotype |
| *Rap1gap* | increased red blood cell distribution width | hematopoietic system phenotype |

| | | |
|---|---|---|
| *Rap1gap* | increased circulating amylase level | homeostasis/metabolism phenotype |
| *Rap1gap* | decreased lymphocyte cell number | hematopoietic system phenotype,immune system phenotype |
| *Rap1gap* | increased neutrophil cell number | hematopoietic system phenotype,immune system phenotype |
| *Rap1gap* | increased eosinophil cell number | hematopoietic system phenotype,immune system phenotype |
| *Cpne5* | abnormal spleen morphology | hematopoietic system phenotype,immune system phenotype |
| *Cpne5* | decreased anxiety-related response | behavior/neurological phenotype |
| *Mymx* | primary atelectasis | respiratory system phenotype |
| *Mymx* | abnormal muscle morphology | muscle phenotype |
| *Mymx* | abnormal limb morphology | limbs/digits/tail phenotype |
| *Mymx* | abnormal myoblast fusion | muscle phenotype,cellular phenotype |
| *Mymx* | muscle weakness | muscle phenotype |
| *Mymx* | cyanosis | homeostasis/metabolism phenotype |
| *Mymx* | abnormal limb muscle morphology | muscle phenotype,limbs/digits/tail phenotype |
| *Mymx* | abnormal spine curvature | skeleton phenotype |
| *Mymx* | abnormal intercostal muscle morphology | muscle phenotype |
| *Mymx* | nuchal edema | homeostasis/metabolism phenotype,integument phenotype,growth/size/body region phenotype |
| *Mymx* | abnormal skeletal muscle morphology | muscle phenotype |
| *Mymx* | decreased fetal weight | growth/size/body region phenotype |
| *Mymx* | neonatal lethality, complete penetrance | mortality/aging |
| *Yy1* | no abnormal phenotype detected | normal phenotype |
| *Yy1* | cyanosis | homeostasis/metabolism phenotype |
| *Yy1* | decreased cell proliferation | cellular phenotype |
| *Yy1* | prenatal lethality, incomplete penetrance | mortality/aging |
| *Yy1* | respiratory failure | respiratory system phenotype |
| *Yy1* | exencephaly | nervous system phenotype |
| *Yy1* | decreased birth body size | growth/size/body region phenotype |
| *Yy1* | decreased embryo size | embryo phenotype,growth/size/body region phenotype |
| *Yy1* | lethality throughout fetal growth and development, incomplete penetrance | mortality/aging |
| *Yy1* | absent egg cylinders | embryo phenotype |
| *Yy1* | pallor | integument phenotype |
| *Yy1* | neonatal lethality, complete penetrance | mortality/aging |
| *Yy1* | embryonic lethality, complete penetrance | mortality/aging |
| *Iqcb1* | decreased total body fat amount | adipose tissue phenotype |
| *Iqcb1* | decreased thigmotaxis | behavior/neurological phenotype |
| *Iqcb1* | increased lean body mass | growth/size/body region phenotype |

| | | |
|---|---|---|
| *Iqcb1* | increased cornea thickness | vision/eye phenotype |
| *Iqcb1* | no abnormal phenotype detected | normal phenotype |
| *Iqcb1* | thick ventricular wall | cardiovascular system phenotype |
| *Iqcb1* | preweaning lethality, incomplete penetrance | mortality/aging |
| *Iqcb1* | increased heart left ventricle size | cardiovascular system phenotype |
| *Coro1a* | enlarged lymph nodes | immune system phenotype |
| *Coro1a* | decreased spleen weight | hematopoietic system phenotype,immune system phenotype |
| *Coro1a* | increased spleen weight | hematopoietic system phenotype,immune system phenotype,growth/size/body region phenotype |
| *Coro1a* | no abnormal phenotype detected | normal phenotype |
| *Coro1a* | abnormal T cell activation | hematopoietic system phenotype,immune system phenotype |
| *Coro1a* | abnormal T cell physiology | hematopoietic system phenotype,immune system phenotype |
| *Coro1a* | decreased susceptibility to bacterial infection | immune system phenotype |
| *Coro1a* | decreased susceptibility to systemic lupus erythematosus | immune system phenotype |
| *Coro1a* | abnormal thymocyte activation | hematopoietic system phenotype,immune system phenotype,endocrine/exocrine gland phenotype |
| *Coro1a* | decreased memory T cell number | hematopoietic system phenotype,immune system phenotype |
| *Coro1a* | increased mast cell degranulation | hematopoietic system phenotype,immune system phenotype,cellular phenotype |
| *Coro1a* | decreased thymocyte number | hematopoietic system phenotype,immune system phenotype,endocrine/exocrine gland phenotype |
| *Coro1a* | glomerulonephritis | immune system phenotype,renal/urinary system phenotype |
| *Coro1a* | decreased T cell number | hematopoietic system phenotype,immune system phenotype |
| *Coro1a* | increased thymocyte number | hematopoietic system phenotype,immune system phenotype,endocrine/exocrine gland phenotype |
| *Coro1a* | small lymph nodes | immune system phenotype |
| *Coro1a* | decreased autoantibody level | immune system phenotype |
| *Coro1a* | abnormal effector T cell morphology | hematopoietic system phenotype,immune system phenotype |
| *Coro1a* | decreased interleukin-6 secretion | immune system phenotype |
| *Coro1a* | increased T cell apoptosis | hematopoietic system phenotype,immune system phenotype,cellular phenotype |
| *Coro1a* | impaired macrophage phagocytosis | hematopoietic system phenotype,immune system phenotype,cellular phenotype |
| *Coro1a* | abnormal leukocyte migration | hematopoietic system phenotype,immune system phenotype,cellular phenotype |
| *Coro1a* | hyperactivity | behavior/neurological phenotype |
| *Coro1a* | decreased tumor necrosis factor secretion | immune system phenotype |
| *Coro1a* | increased susceptibility to systemic lupus erythematosus | immune system phenotype |

| | | |
|---|---|---|
| *Coro1a* | decreased monocyte cell number | hematopoietic system phenotype,immune system phenotype |
| *Coro1a* | decreased B cell number | hematopoietic system phenotype,immune system phenotype |
| *Coro1a* | decreased T cell proliferation | hematopoietic system phenotype,immune system phenotype,cellular phenotype |
| *Coro1a* | enlarged spleen | hematopoietic system phenotype,immune system phenotype,growth/size/body region phenotype |
| *Coro1a* | decreased CD8-positive, alpha-beta T cell number | hematopoietic system phenotype,immune system phenotype |
| *Coro1a* | decreased interleukin-2 secretion | immune system phenotype |
| *Coro1a* | decreased CD4-positive, alpha-beta T cell number | hematopoietic system phenotype,immune system phenotype |
| *Cpsf4l* | increased circulating alanine transaminase level | homeostasis/metabolism phenotype |
| *Wdr19* | absent frontal bone | craniofacial phenotype,skeleton phenotype |
| *Wdr19* | abnormal spine curvature | skeleton phenotype |
| *Wdr19* | split sternum | skeleton phenotype |
| *Wdr19* | abnormal premaxilla morphology | craniofacial phenotype,skeleton phenotype |
| *Wdr19* | abnormal limb paddle morphology | limbs/digits/tail phenotype |
| *Wdr19* | abnormal maxilla morphology | craniofacial phenotype,skeleton phenotype |
| *Wdr19* | abnormal cilium morphology | cellular phenotype |
| *Wdr19* | abnormal craniofacial development | craniofacial phenotype |
| *Wdr19* | short face | craniofacial phenotype,growth/size/body region phenotype |
| *Wdr19* | abnormal tympanic ring morphology | hearing/vestibular/ear phenotype |
| *Wdr19* | interparietal bone hypoplasia | craniofacial phenotype,skeleton phenotype |
| *Wdr19* | cleft palate | craniofacial phenotype,digestive/alimentary phenotype,growth/size/body region phenotype |
| *Wdr19* | embryonic lethality during organogenesis, complete penetrance | mortality/aging |
| *Wdr19* | short ribs | skeleton phenotype |
| *Wdr19* | absent supraoccipital bone | craniofacial phenotype,skeleton phenotype |
| *Wdr19* | abnormal vertebrae morphology | skeleton phenotype |
| *Wdr19* | abnormal mandibular angle morphology | craniofacial phenotype,skeleton phenotype |
| *Wdr19* | polyphalangy | limbs/digits/tail phenotype,skeleton phenotype |
| *Wdr19* | short maxilla | craniofacial phenotype,skeleton phenotype |
| *Wdr19* | abnormal sternum ossification | skeleton phenotype |
| *Wdr19* | polydactyly | limbs/digits/tail phenotype |
| *Wdr19* | absent tibia | limbs/digits/tail phenotype,skeleton phenotype |
| *Wdr19* | abnormal limb mesenchyme morphology | embryo phenotype,limbs/digits/tail phenotype |
| *Wdr19* | abnormal limb development | limbs/digits/tail phenotype |
| *Wdr19* | anophthalmia | vision/eye phenotype |
| *Wdr19* | cleft secondary palate | craniofacial phenotype,digestive/alimentary phenotype,growth/size/body region phenotype |

| | | |
|---|---|---|
| *Wdr19* | abnormal somite development | embryo phenotype |
| *Wdr19* | abnormal rib morphology | skeleton phenotype |
| *Wdr19* | lethality throughout fetal growth and development, complete penetrance | mortality/aging |
| *Wdr19* | rib bifurcation | skeleton phenotype |
| *Wdr19* | ectopic digits | limbs/digits/tail phenotype |
| *Wdr19* | abnormal dermomyotome development | muscle phenotype |
| *Wdr19* | parietal bone hypoplasia | craniofacial phenotype,skeleton phenotype |
| *Wdr19* | abnormal palatal shelf fusion at midline | craniofacial phenotype,digestive/alimentary phenotype,growth/size/body region phenotype |
| *Wdr19* | decreased palatine bone horizontal plate size | craniofacial phenotype,digestive/alimentary phenotype,growth/size/body region phenotype,skeleton phenotype |
| *Wdr19* | short mandible | craniofacial phenotype,skeleton phenotype |
| *Wdr19* | abnormal myotome development | muscle phenotype |
| *Wdr19* | abnormal cranium morphology | craniofacial phenotype,skeleton phenotype |
| *Wdr19* | exencephaly | nervous system phenotype |
| *Wdr19* | rib fusion | skeleton phenotype |
| *Wdr19* | abnormal palatine bone morphology | craniofacial phenotype,skeleton phenotype |
| *Wdr19* | bilateral cleft upper lip | craniofacial phenotype,growth/size/body region phenotype |
| *Wdr19* | short premaxilla | craniofacial phenotype,skeleton phenotype |

| Supplementary Table 13. STRINGdb Analysis Results* | | | | | | |
|---|---|---|---|---|---|---|
| **Category** | **term ID** | **term description** | **background gene count** | **strength** | **false discovery rate** | **matching proteins in your network (labels)** |
| GO Function | GO:0001601 | Peptide YY receptor activity | 3 | 2.91 | 0.0304 | NPY1R,NPY5R |
| GO Function | GO:0001602 | Pancreatic polypeptide receptor activity | 4 | 2.79 | 0.0304 | NPY1R,NPY5R |
| GO Function | GO:0030976 | Thiamine pyrophosphate binding | 8 | 2.49 | 0.0455 | DHTKD1,TKTL2 |
| UniProt Keywords | KW-0786 | Thiamine pyrophosphate | 13 | 2.28 | 0.0433 | DHTKD1,TKTL2 |
| SMART | SM00861 | Transketolase, pyrimidine binding domain | 7 | 2.55 | 0.0182 | DHTKD1,TKTL2 |

*Enriched terms for the protein-protein interaction network identified by STRINGdb when querying whole-genome sequence common variant analysis interaction gene loci (rs281851, rs13118183) reported in Table 1

**Supplementary Table 14. DGIdb Query Results**

| *Gene* | *Drug* | *Interaction Score* | *Interaction Types* | *Sources* | *PMIDs* |
|---|---|---|---|---|---|
| ***CAMK1D*** | LOSARTAN | 1.72 | | PharmGKB | 25410890 |
| ***NPY1R*** | CHEMBL1585652 | 2.58 | | DTC | |
| ***NPY1R*** | CHEMBL422942 | 2.58 | | TTD | |
| ***NPY1R*** | N6-PHENYLADENOSINE | 1.1 | | DTC | |
| ***NPY1R*** | CHEMBL530291 | 1.1 | | DTC | |
| ***NPY1R*** | NEUROPEPTIDE-Y | 3.86 | | TTD | |
| ***NPY1R*** | CHEMBL324554 | 7.73 | | DTC | |
| ***NPY5R*** | VELNEPERIT | 30.91 | antagonist | TdgClinicalTrial\|ChemblInteractions | |
| ***NPY5R*** | S-237648 | 15.46 | antagonist | ChemblInteractions | |
| ***NPY5R*** | PEPTIDE YY HUMAN (3-36) | 5.15 | | TdgClinicalTrial | |
| ***NPY5R*** | FR-79620 | 15.46 | | TTD | |
| ***EYA2*** | Solypertine Tartrate | 1.03 | | DTC | |
| ***EYA2*** | (R)-Edelfosine | 1.03 | | DTC | |
| ***EYA2*** | Hexamethyl Pararosaniline | 1.03 | | DTC | |

**Supplementary Table 15. QIAGEN Canonical Pathway Enrichment Analysis Results**

| Genes | Pathway | P-Value |
|---|---|---|
| ***PI4K2B, UBLCP1*** | 3-phosphoinositide Biosynthesis | **8.13E-03** |
| ***PI4K2B, UBLCP1*** | Superpathway of Inositol Phosphate Compounds | **1.05E-02** |
| ***RAP1GAP*** | Rap1 signalling | **1.05E-02** |
| ***PI4K2B*** | D-myo-inositol (1,4,5)-Trisphosphate Biosynthesis | **1.70E-02** |
| ***YY1*** | Transcriptional regulation by the AP-2 (TFAP2) family of transcription factors | **2.34E-02** |
| ***RAP1GAP*** | RET signaling | **2.69E-02** |
| ***MED31, ZNF773*** | Generic Transcription Pathway | **3.16E-02** |
| ***SCUBE2*** | Hedgehog ligand biogenesis | **4.17E-02** |
| ***YY1*** | VDR/RXR Activation | 5.01E-02 |
| ***YY1*** | Activation of anterior HOX genes in hindbrain during early embryogenesis | 5.25E-02 |
| ***PI4K2B*** | PI Metabolism | 5.25E-02 |
| ***MED31*** | Transcriptional regulation of white adipocyte differentiation | 5.37E-02 |
| ***YY1*** | ESR-mediated signaling | 7.59E-02 |
| ***MED31*** | Regulation of lipid metabolism by PPARalpha | 7.59E-02 |
| ***RAP1GAP*** | Endocannabinoid Developing Neuron Pathway | 8.13E-02 |
| ***RAP1GAP*** | Gai Signaling | 8.91E-02 |
