## Supplementary figures and images for "Gene-Excessive Sleepiness Interactions Suggest Treatment Targets for Obstructive Sleep Apnea Subtype"

### Supplementary Figures

Supplementary Figure 1. QIAGEN Ingenuity Pathway Analysis Results

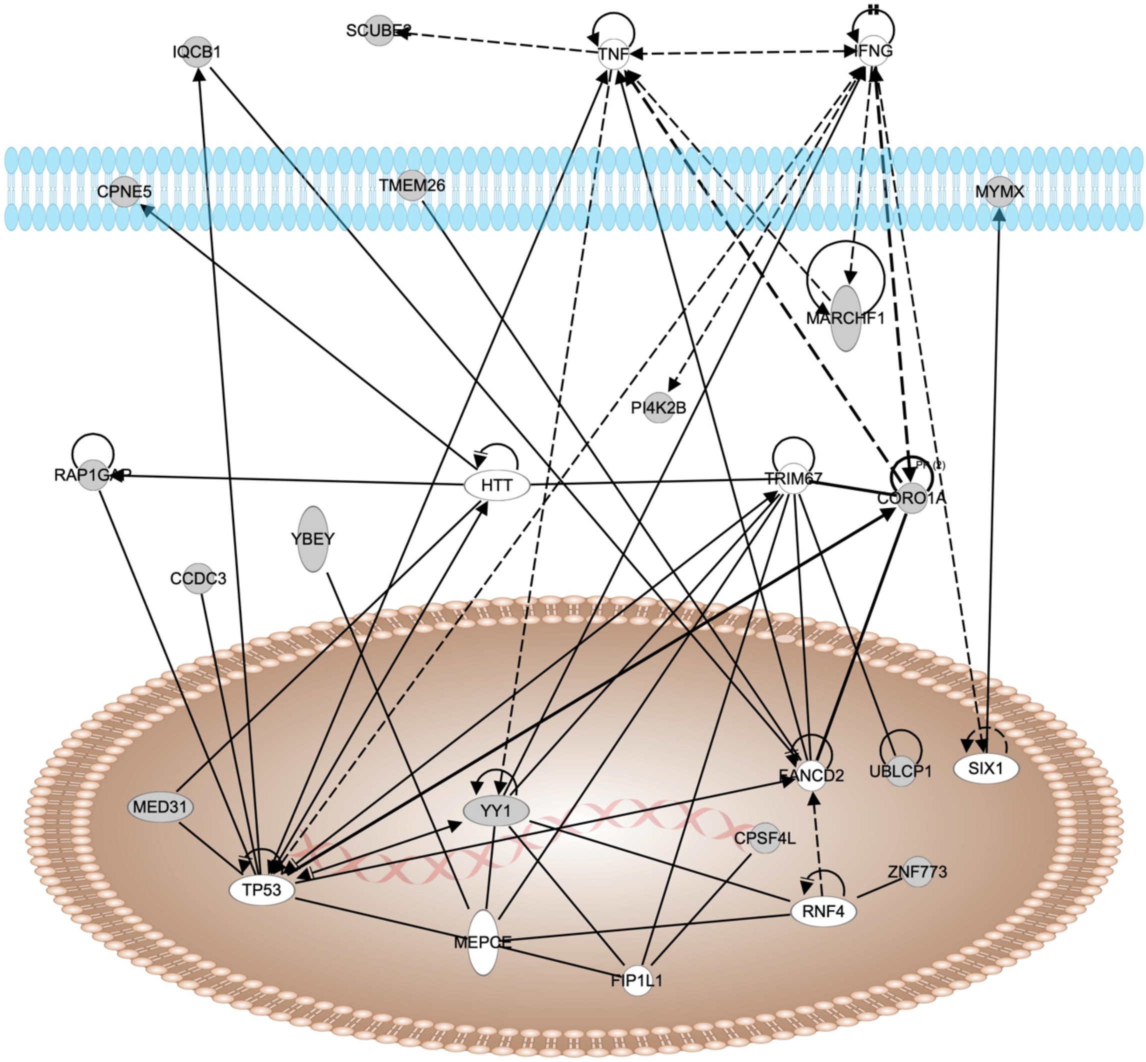


© 2000-2024 QIAGEN. All rights reserved.
