## Supplementary Note for "Gene-Excessive Sleepiness Interactions Suggest Treatment Targets for Obstructive Sleep Apnea Subtype"

### COHORT DESCRIPTIONS

#### Atherosclerosis Risk in Communities (ARIC)

ARIC is a prospective longitudinal study initiated in 1985, with community surveillance performed for cardiovascular disease between 1987 to 2014, and 33 years of follow-up to identify health events in an established cohort of African Americans and white individuals [1]. Sleep measures were collected as a part of the Sleep Heart Health Study (SHHS) conducted in 1995 to 1998 [2]. Detailed descriptions of SHHS can be found in the National Sleep Research Resource [3] and of ARIC at the database of Genotypes and Phenotypes (dbGaP) with accession numbers phs001211 and phs000280.

#### Cardiovascular Health Study (CHS)

CHS is a prospective longitudinal cohort that was initiated in 1989 and was designed to investigate cardiovascular measures including disease risk factors, subclinical disease measures, occurrence of myocardial infarction and stroke, and venous thromboembolism [4]. Sleep measures were collected as a part of the Sleep Heart Health Study (SHHS) conducted in 1995 to 1998 [2]. Detailed descriptions of SHHS can be found in the National Sleep Research Resource [3] and of CHS at dbGaP with accession numbers phs001368 and phs000287.

#### Framingham Heart Study (FHS)

FHS is a prospective longitudinal study that began in 1948, to study atherosclerosis development and further clinical outcomes. Sleep measures were collected as a part of the Sleep Heart Health Study (SHHS) conducted in 1995 to 1998 [2]. Detailed descriptions of SHHS can be found in the National Sleep Research Resource [3] and of FHS at dbGaP with accession numbers phs000974 and phs000007.

#### Cleveland Family Study (CFS)

CFS is a prospective longitudinal family-based study that was conducted between 1990 to 2006 for 261 families, designed to identify probands diagnosed with obstructive sleep apnea. Detailed descriptions of CFS can be found at dbGaP with accession numbers phs000954 and phs000284, and the National Sleep Research Resource [3,5,6].

#### Hispanic Community Health Study/Study of Latinos (HCHS/SOL)

HCHS/SOL is a prospective longitudinal study conducted across multiple centers for individuals of Cuban, Dominican, Mexican, Puerto Rican, Central American, and South American backgrounds. Detailed descriptions of HCHS/SOL can be found in the National Sleep Research Resource [3] and at dbGaP with accession numbers phs001395 and phs000810.

#### Jackson Heart Study (JHS)

JHS is a prospective longitudinal cohort formed to investigate cardiovascular disease among African Americans residing in urban and rural areas of Jackson, MS, metropolitan statistical area

[7]. Detailed descriptions of JHS can be found at dbGaP with accession numbers phs000964 and phs000286.

#### Multi-Ethnic Study of Atherosclerosis (MESA)

MESA is a prospective longitudinal study of a set of diverse 6814 individuals conducted across 6 field centers, focused on assessment of subclinical cardiovascular disease and risk factors for progression to clinically overt cardiovascular disease or progression of subclinical disease [8]. A subset of individuals were enrolled in MESA Sleep in 2010-2012 which incorporated polysomnography, actigraphy, and sleep questionnaire measurements. Detailed descriptions can be found in the National Sleep Research Resource [3] and at dbGaP with accession numbers phs001416 and phs000209.

#### The Osteoporotic Fractures in Men Study (MrOS)

The Osteoporotic Fractures in Men Study's ancillary sleep study (population-based, cross-sectional) is defined as MrOS, consisting of individuals who underwent polysomnography and actigraphy studies between December 2003 to March 2005 [9]. A study description of MrOS can be found in the National Sleep Research Resource, and MrOS design and recruitment publications [3,10,11].

#### Western Australian Sleep Health Study (WASHS)

WASHS is a study of 4000 patients with obstructive sleep apnea from a public sleep clinic in Western Australia, with questionnaire, biochemistry, DNA, and polysomnography data collected between 2006-2010 [12].

### DATA CONSOLIDATION

In CFS, SHHS, MESA, MrOS, and WASHS, apnea hypopnea index (AHI) was defined as the number of episodes of complete (apnea) or partial (hypopnea) cessations of airflow per hour of sleep, with a minimum desaturation of 3%. Apart from WASHS, all sleep data were scored by blinded scorers from the same central Sleep Reading Center with high levels of inter- and intra-scorer reliability using well-defined procedures [13,14]. Individuals who self-identified as European descent were retained for MrOS and WASHS cohorts.

### REFERENCES


1. Wright, J.D. *et al.* The ARIC (Atherosclerosis Risk In Communities) Study: JACC Focus Seminar 3/8. *J Am Coll Cardiol* **77**, 2939-2959 (2021).
2. Quan, S.F. *et al.* The Sleep Heart Health Study: design, rationale, and methods. *Sleep* **20**, 1077-85 (1997).
3. Zhang, G.Q. *et al.* The National Sleep Research Resource: towards a sleep data commons. *J Am Med Inform Assoc* **25**, 1351-1358 (2018).
4. Fried, L.P. *et al.* The Cardiovascular Health Study: design and rationale. *Ann Epidemiol* **1**, 263-76 (1991).
5. Redline, S. *et al.* The familial aggregation of obstructive sleep apnea. *Am J Respir Crit Care Med* **151**, 682-7 (1995).
6. Redline, S., Schluchter, M.D., Larkin, E.K. & Tishler, P.V. Predictors of longitudinal change in sleep-disordered breathing in a nonclinic population. *Sleep* **26**, 703-9 (2003).
7. Wyatt, S.B. *et al.* A community-driven model of research participation: the Jackson Heart Study Participant Recruitment and Retention Study. *Ethn Dis* **13**, 438-55 (2003).
8. Bild, D.E. *et al.* Multi-Ethnic Study of Atherosclerosis: objectives and design. *Am J Epidemiol* **156**, 871-81 (2002).
9. Blackwell, T. *et al.* Associations between sleep architecture and sleep-disordered breathing and cognition in older community-dwelling men: the Osteoporotic Fractures in Men Sleep Study. *J Am Geriatr Soc* **59**, 2217-25 (2011).
10. Orwoll, E. *et al.* Design and baseline characteristics of the osteoporotic fractures in men (MrOS) study--a large observational study of the determinants of fracture in older men. *Contemp Clin Trials* **26**, 569-85 (2005).
11. Blank, J.B. *et al.* Overview of recruitment for the osteoporotic fractures in men study (MrOS). *Contemp Clin Trials* **26**, 557-68 (2005).
12. Mukherjee, S. *et al.* Cohort profile: the Western Australian Sleep Health Study. *Sleep Breath* **16**, 205-15 (2012).
13. Redline, S. *et al.* Methods for obtaining and analyzing unattended polysomnography data for a multicenter study. Sleep Heart Health Research Group. *Sleep* **21**, 759-67 (1998).
14. Whitney, C.W. *et al.* Reliability of scoring respiratory disturbance indices and sleep staging. *Sleep* **21**, 749-57 (1998).
